# Strong humoral immune responses against SARS-CoV-2 Spike after BNT162b2 mRNA vaccination with a 16-week interval between doses

**DOI:** 10.1101/2021.09.17.21263532

**Authors:** Alexandra Tauzin, Shang Yu Gong, Guillaume Beaudoin-Bussières, Dani Vézina, Romain Gasser, Lauriane Nault, Lorie Marchitto, Mehdi Benlarbi, Debashree Chatterjee, Manon Nayrac, Annemarie Laumaea, Jérémie Prévost, Marianne Boutin, Gérémy Sannier, Alexandre Nicolas, Catherine Bourassa, Gabrielle Gendron-Lepage, Halima Medjahed, Guillaume Goyette, Yuxia Bo, Josée Perreault, Laurie Gokool, Chantal Morrisseau, Pascale Arlotto, Renée Bazin, Mathieu Dubé, Gaston De Serres, Nicholas Brousseau, Jonathan Richard, Roberta Rovito, Marceline Côté, Cécile Tremblay, Giulia C. Marchetti, Ralf Duerr, Valérie Martel-Laferrière, Daniel E. Kaufmann, Andrés Finzi

## Abstract

While the standard regimen of the BNT162b2 mRNA vaccine includes two doses administered three weeks apart, some public health authorities decided to space them, raising concerns about vaccine efficacy. Here, we analyzed longitudinal humoral responses including antibody binding, Fc-mediated effector functions and neutralizing activity against the D614G strain but also variants of concern and SARS-CoV-1 in a cohort of SARS-CoV-2 naïve and previously infected individuals, with an interval of sixteen weeks between the two doses. While the administration of a second dose to previously infected individuals did not significantly improve humoral responses, we observed a significant increase of humoral responses in naïve individuals after the 16-weeks delayed second shot, achieving similar levels as in previously infected individuals. We compared these responses to those elicited in individuals receiving a short (4-weeks) dose interval. For the naïve donors, these responses were superior to those elicited by the short dose interval.

## INTRODUCTION

Since the end of 2019, the etiological agent of the Coronavirus disease 2019 (COVID-19), the Severe Acute Respiratory Syndrome Coronavirus-2 (SARS-CoV-2) has spread worldwide causing the current pandemic (Dong et al., 2020; World Health Organization). In the last months, several vaccines against SARS-CoV-2 have been approved in many countries, including the Pfizer/BioNtech BNT162b2 mRNA vaccine. This vaccine targets the highly immunogenic trimeric Spike (S) glycoprotein that facilitates SARS-CoV-2 entry into host cells via its receptor-binding domain (RBD) that interacts with angiotensin-converting enzyme 2 (ACE-2) (Hoffmann et al., 2020; Walls et al., 2020) and has shown an important vaccine efficacy (Polack et al., 2020; Skowronski and De Serres, 2021).

The approved BNT162b2 mRNA vaccine regimen comprises two doses administered 3-4 weeks apart (WHO, 2021). However, at the beginning of the vaccination campaign (Winter/Spring 2021) vaccine scarcity prompted some public health agencies to extend the interval between doses in order to maximize the number of immunized individuals. This strategy was supported by results indicating that a single dose affords ∼90% protection starting two weeks post vaccination, concomitant with the detection of some vaccine-elicited immune responses (Baden et al., 2021; Pilishvili, 2021; Polack et al., 2020; Skowronski and De Serres, 2021; Tauzin et al., 2021).

The rapid emergence of several variants of concerns (VOCs) and variants of interest (VOIs), which are more transmissible and in some cases more virulent (Allen et al., 2021; Brown et al., 2021; Davies et al., 2021; Fisman and Tuite, 2021; Pearson et al., 2021) remains a major public health preoccupation as the vaccine campaign advances worldwide. For example, the mutation D614G in the S glycoprotein which appeared very early in the pandemic is now present in almost all circulating strains (Isabel et al., 2020). The B.1.1.7 (Alpha) variant emerged in late 2020 in the United Kingdom and due to its increased affinity for the ACE2 receptor that leads to increased transmissibility (Davies et al., 2021), it became in just a few months a predominant strain worldwide (Davies et al., 2021; Prévost et al., 2021; Rambaut et al., 2020). The B.1.351 (Beta) and P.1 (Gamma) variants that first emerged in South Africa and Brazil respectively have largely spread and are now circulating in many countries (ECDC, 2021; Tang et al., 2021). The B.1.526 (Iota) variant first identified in New York in early 2021 is in an upward trajectory in the United States (Annavajhala et al., 2021). More recently, the B.1.617.2 (Delta) variant which emerged in India and has a high transmissibility is now the dominant strain in several countries (Allen et al., 2021; Dagpunar, 2021). Although several studies have shown that mRNA vaccines protect against severe disease caused by these variants, it has also been shown that some of them present resistance to some vaccine-elicited immune responses, notably against neutralizing antibodies (Annavajhala et al., 2021; Goel et al., 2021a; Planas et al., 2021a; Puranik et al., 2021; Wall et al., 2021; Wang et al., 2021a). Most of these studies were based on the analysis of plasma samples collected from vaccinees following a short (3-4 weeks) interval between doses. Little is known about vaccine-elicited immune responses with longer dose intervals. Here, we characterized vaccine-elicited humoral responses in a cohort of SARS-CoV-2 naïve and previously infected individuals that received the two doses with an extended interval of sixteen weeks.

## RESULTS

We analyzed the longitudinal humoral responses after vaccination with the BNT162b2 mRNA vaccine in blood samples, with an interval of around 16 weeks between the two doses (median [range]: 111 days [76–134 days]). The cohort included 26 SARS-CoV-2 naïve and 27 previously infected (PI) donors tested SARS-CoV-2 positive by nasopharyngeal swab PCR around 9 months before their first dose (median [range]: 281 days [116-342 days]). In the cohort of PI individuals, 12 donors did not receive the second injection, leaving 15 PI donors with two doses. The blood samples were collected at different time points: prior the first dose of vaccine (V0), three weeks (V1, median [range]: 20 days [13–28 days]) and three months (V2, median [range]: 84 days [67–104 days]) after the first dose of vaccine, and three weeks (V3, median [range]: 22 days [13–51 days]) and four months (V4, median [range]: 113 days [90-127 days]) after the second vaccine injection. Data collected at V0 and V1 have been previously described (Tauzin et al., 2021). Basic demographic characteristics of the cohorts and detailed vaccination timepoints are summarized in Table 1 and Figure 1A.

**Figure 1.**
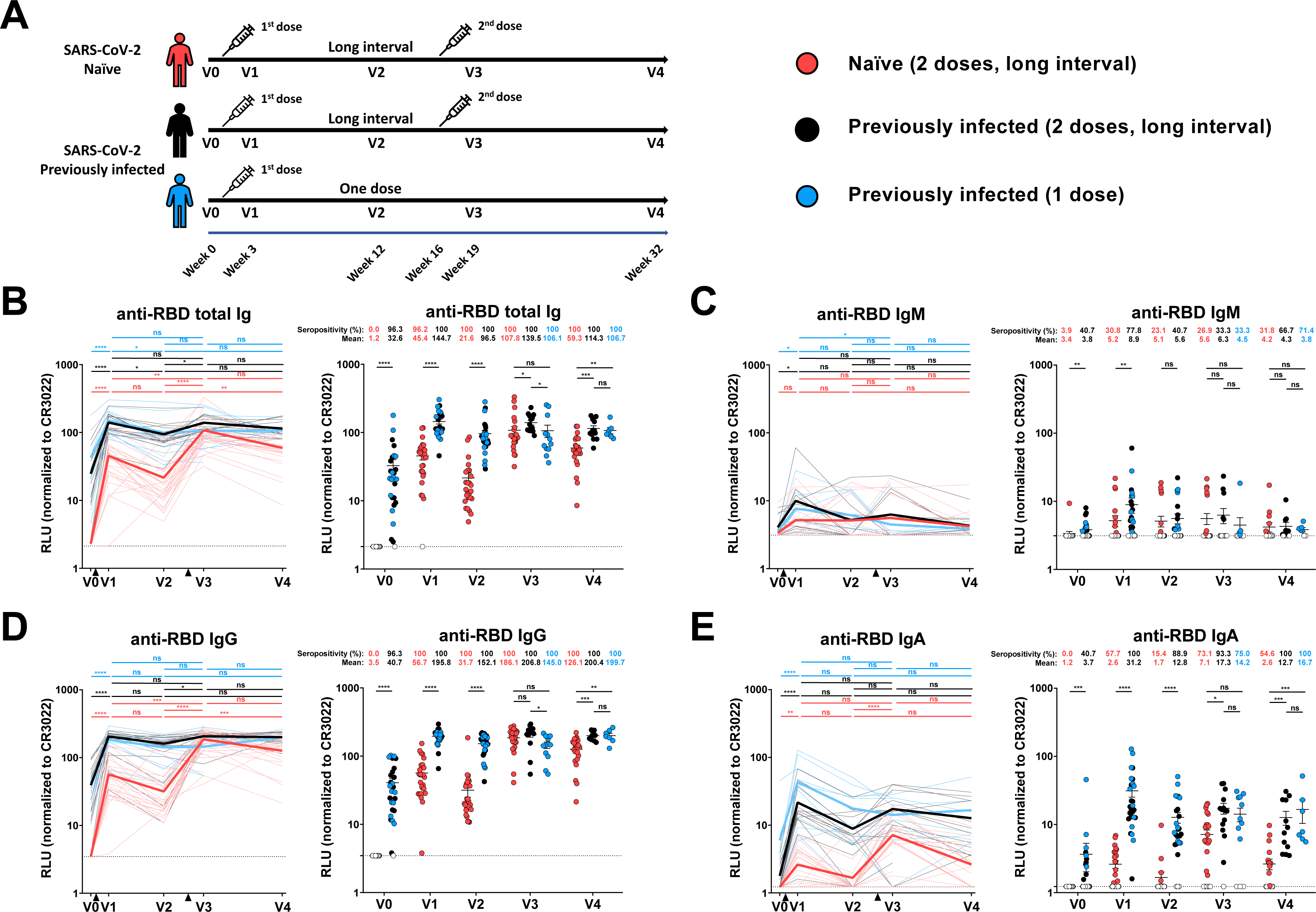
Elicitation of RBD-specific antibodies in SARS-CoV-2 naïve and previously-infected individuals. (**A**) SARS-CoV-2 vaccine cohort design. (**B-E**) Indirect ELISA was performed by incubating plasma samples from naïve and PI donors collected at V0, V1, V2, V3 and V4 with recombinant SARS-CoV-2 RBD protein. Anti-RBD Ab binding was detected using HRP-conjugated (**B**) anti-human IgM+IgG+IgA (**C**) anti-human IgM, (**D**) anti-human IgG, or (**E**) anti-human IgA. Relative light unit (RLU) values obtained with BSA (negative control) were subtracted and further normalized to the signal obtained with the anti-RBD CR3022 mAb present in each plate. Naïve and PI donors with a long interval between the two doses are represented by red and black points respectively and PI donors who received just one dose by blue points. (**Left panels**) Each curve represents the normalized RLUs obtained with the plasma of one donor at every time point. Mean of each group is represented by a bold line. The time of vaccine dose injections is indicated by black triangles. (**Right panels**) Plasma samples were grouped in different time points (V0, V1, V2, V3 and V4). Undetectable measures are represented as white symbols, and limits of detection are plotted. Error bars indicate means ± SEM. (* P < 0.05; ** P < 0.01; *** P < 0.001; **** P < 0.0001; ns, non-significant).

**Table 1.** Characteristics of the vaccinated SARS-CoV-2 cohorts.

|  |  | SARS-CoV-2 Naïve |  | SARS-CoV-2 Previously infected |  |  |
| --- | --- | --- | --- | --- | --- | --- |
|  |  | Two doses<br>Short interval<br>(n=12) | Two doses<br>Long interval<br>(n=26) | Two doses<br>Long interval<br>(n=15) | One dose<br>(n=12) | Entire cohort<br>(n=27) |
| Age |  | 45 (24-60) | 50 (21-62) | 47 (29-65) | 51 (21-65) | 48 (21-65) |
| Gender | Male (n) | 8 | 11 | 10 | 4 | 14 |
|  | Female (n) | 4 | 15 | 5 | 8 | 13 |
| Days between symptom onset and the 1<sup>st</sup> dose <sup>a</sup> |  | N/A | N/A | 274 (166-321) | 288 (116-342) | 281 (116-342) |
| Days between the 1<sup>st</sup> and 2<sup>nd</sup> dose <sup>a</sup> |  | 30 (22-34) | 111 (76-120) | 110 (90-134) | N/A | N/A |
| Days between V0 and the 1<sup>st</sup> dose <sup>a</sup> |  | N/A | 1 (0-86) | 24 (0-95) | 18 (1-117) | 23 (0-117) |
| Days between the 1<sup>st</sup> dose and V1 <sup>a</sup> |  | N/A | 21 (16-28) | 20 (17-25) | 20 (13-21) | 20 (13-25) |
| Days between the 1<sup>st</sup> dose and V2 <sup>a</sup> |  | N/A | 83 (67-92) | 89 (82-104) | 90 (80-104) | 89 (80-104) |
| Days between V2 and the 2<sup>nd</sup> dose <sup>a</sup> |  | N/A | 28 (9-38) | 23 (2-42) | N/A | N/A |
| Days between the 1<sup>st</sup> dose and V3 <sup>a</sup> |  | 54 (41-65) | 133 (102-144) | 138 (103-161) | 132 (120-146) | 136 (103-161) |
| Days between the 2<sup>nd</sup> dose and V3 <sup>a</sup> |  | 24 (12-37) | 21 (14-34) | 22 (13-51) | N/A | N/A |
| Days between the 1<sup>st</sup> dose and V4 <sup>a</sup> |  | N/A | 224 (215-237) | 225 (215-248) | 231 (222-248) | 225 (215-248) |
| Days between the 2<sup>nd</sup> dose and V4 <sup>a</sup> |  | N/A | 112 (103-125) | 113 (90-127) | N/A | N/A |

<sup>a</sup> Values displayed are medians, with ranges in parentheses.

### Elicitation of SARS-CoV-2 antibodies against the full Spike and its receptor-binding domain

To evaluate vaccine responses in SARS-CoV-2 naïve and PI individuals, we first measured the presence of SARS-CoV-2-specific antibodies (Abs) (IgG, IgM, IgA) recognizing the receptor-binding domain (Figure 1B-E) using an ELISA RBD assay or the native full-length S glycoprotein expressed at the cell surface (Figure S1A-D) using a cell-based ELISA assay. Both assays have been previously described (Anand et al., 2021; Beaudoin-Bussières et al., 2020; Prévost et al., 2020). Prior to vaccination (V0), no SARS-CoV-2 specific Abs were detectable in SARS-CoV-2 naïve individuals, except for anti-Spike IgM (26.9% seropositivity) which are likely to be cross-reactive antibodies against the S2 subunit (Fraley et al., 2021; Hicks et al., 2021; Ng et al., 2020). SARS-CoV-2 PI individuals still had detectable Abs several months post-symptoms onset, especially IgG, in agreement with previous observations (Anand et al., 2021; Dan et al., 2021; Tauzin et al., 2021; Wang et al., 2021b). For both groups, the first dose of vaccine induced a significant increase of total immunoglobulins (Igs) recognizing the RBD or the Spike protein three weeks post-vaccine (V1), with a significantly higher response for the PI group (Figure 1B-E and S1A-D). At V2 (i.e., 12 weeks post vaccination), while anti-Spike total Ig levels remained stable, we observed a decrease in anti-RBD total Ig levels in both groups, with the exception of some naïve donors where we observed an increase. We did not detect Abs recognizing the N protein for these donors (not shown), suggesting that they had not been infected between the two doses. This increase could therefore be linked to a delayed response or affinity maturation of the antibodies in the germinal center between V1 and V2. The second dose, which was administered ∼16 weeks after the first one, strongly boosted the induction of anti-RBD Igs in the SARS-CoV-2 naïve group, particularly IgG and IgA which reached higher levels (Figure 1D and E). For the PI group, the second dose also led to an increase in the level of total anti-RBD Igs similar to that achieved after the first dose. Of note, the second dose in the naïve group elicited anti-RBD IgG levels that reached the same levels than in the PI group receiving one or two doses (Figure 1D). However, four months after the second dose (V4), we observed a decrease in anti-RBD Igs that was more important in the naïve group compared to the PI groups. Also, we noted that PI individuals always had a higher level of anti-RBD IgA than naïve individuals at every time point (Figure 1E). Similar patterns of responses were observed when we measured the level of Abs recognizing the full-length S glycoprotein (Figure S1A-D).

### Recognition of SARS-CoV-2 Spike variants and other *Betacoronaviruses*

The BNT162b2 mRNA vaccine has been developed against the original Wuhan strain. However, SARS-CoV-2 is evolving, and many variants have emerged and spread rapidly worldwide. Some harbor specific mutations in S that are associated with increased transmissibility and/or immune evasion (Davies et al., 2021; Sabino et al., 2021; Tegally et al., 2020; Volz et al., 2021). Here, we evaluated the ability of Abs elicited by the Pfizer/BioNTech vaccine to recognize different S proteins of VOCs (B.1.1.7, B.1.351, P.1 and B.1.612.2) and the VOI B.1.526 expressed at the cell surface of 293T cells by flow cytometry, using a method we have previously described (Figure 2, S2) (Gong et al., 2021; Prévost et al., 2020; Tauzin et al., 2021).

**Figure 2.**
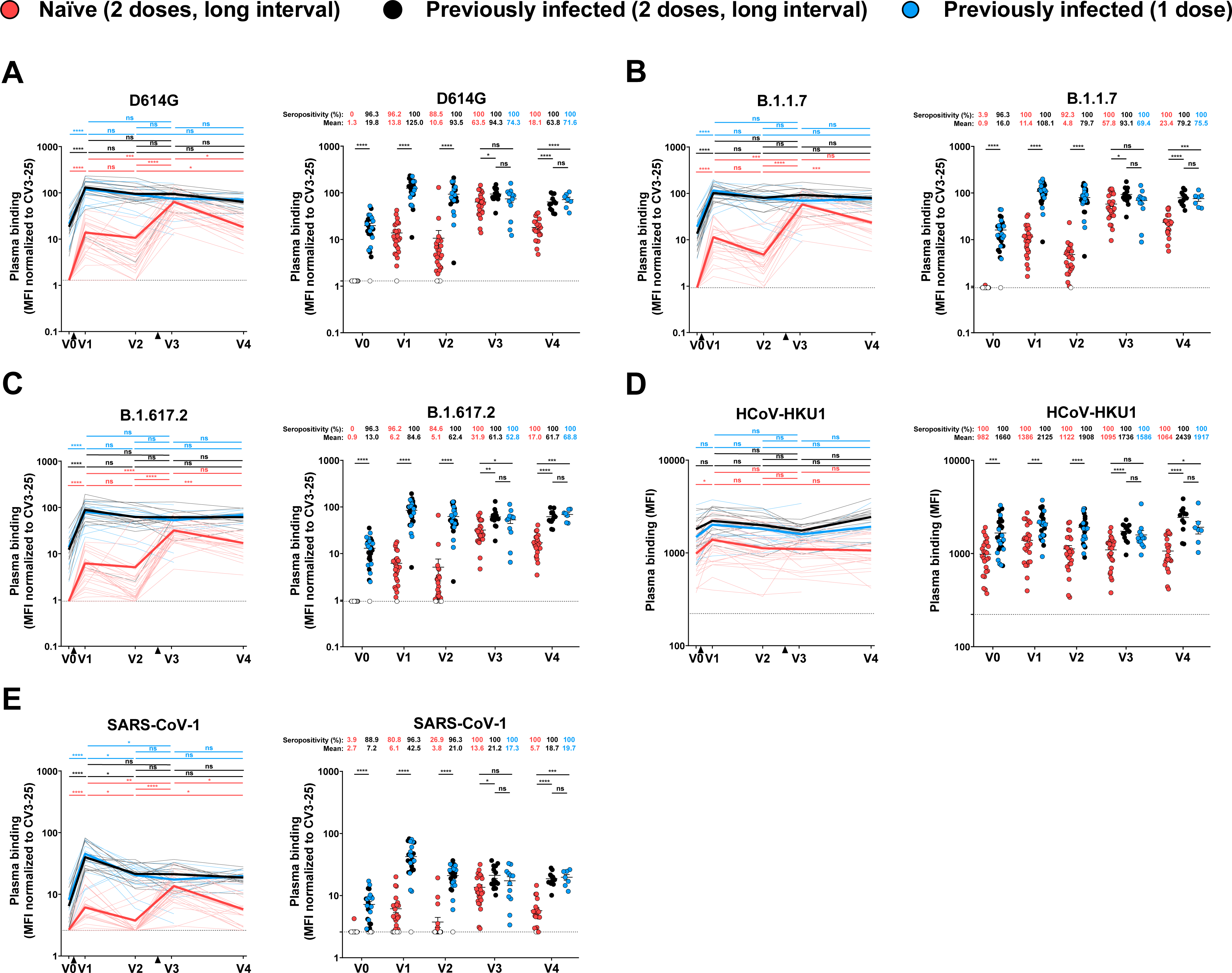
Binding of vaccine-elicited antibodies to SARS-CoV-2 Spike variants and other *Betacoronaviruses*. 293T cells were transfected with the indicated full-length S from different SARS-CoV-2 variants and other human *Betacoronavirus* Spikes and stained with the CV3-25 Ab or with plasma from naïve or PI donors collected at V0, V1, V2, V3 and V4 and analyzed by flow cytometry. The values represent the median fluorescence intensities (MFI) (**D**) or the MFI normalized by CV3-25 Ab binding (**A-C**, **E**). Naïve and PI donors with a long interval between the two doses are represented by red and black points respectively and PI donors who received just one dose by blue points. (**Left panels**) Each curve represents the MFI or the normalized MFIs obtained with the plasma of one donor at every time point. Mean of each group is represented by a bold line. The time of vaccine dose injections is indicated by black triangles. (**Right panels**) Plasma samples were grouped in different time points (V0, V1, V2, V3 and V4). Undetectable measures are represented as white symbols, and limits of detection are plotted. Error bars indicate means ± SEM. (* P < 0.05; ** P < 0.01; *** P < 0.001; **** P < 0.0001; ns, non-significant).

As expected, none of the SARS-CoV-2 naïve plasma samples collected at V0 were able to recognize the SARS-CoV-2 S (D614G) or any of the variants tested here (B.1.1.7, B.1.351, B.1.617.2, P.1, B.1.526) (Figure 2A-C and S2A). In contrast, plasma from PI individuals recognized all tested SARS-CoV-2 variants at V0 (Figure 2A-C, S2A). The first dose of vaccine strongly enhanced the recognition of the full D614G S and all the tested variants in both groups (Figure 2A-C and S2B). Three months after the first dose, the recognition slightly decreased but not significantly. As expected, the second dose strongly increased recognition of all VOC Spikes in the naïve group and reached levels that where significantly higher than after the first dose. In contrast, for the PI group, the second dose did not result in a better recognition than after the first dose. Of note, we observed no significant differences at V3 between PI individuals who received one or two doses, despite a shorter period since the last dose for PI individuals who received two doses. The recognition of all VOCs was slightly lower at V3 by the naïve group compared to the PI that received two doses (Figure 2A-C). When we compared Spike recognition between the SARS-CoV-2 variants, we observed that plasma from PI individuals before vaccination recognized less efficiently the different S variants compared to the D614G S (Figure S2A). After the first and second dose, only B.1.351 and B.1.617.2 S were less efficiently recognized by plasmas from PI individuals (Figure S2B-D). For naïve individuals, even if the vaccination strongly increased the recognition of every VOC Spike tested, we observed that plasmas recognized the different SARS-CoV-2 variants less efficiently compared to D614G S except for the B.1.1.7 S after the second dose (Figure S2). As observed for the level of anti-RBD Igs, (Figure 1), while the recognition of the different SARS-CoV-2 Spikes at V4 (i.e., 4 months after the second dose) remained stable in the PI group, it decreased in the naïve group at V4 (Figure 2A-C).

We also evaluated whether vaccination elicited Abs that were able to recognize S glycoproteins from endemic human *Betacoronaviruses*, (HCoV-HKU1). Interestingly, we observed that the first but not the second dose enhanced the recognition of HCoV-HKU1 S in the naïve group (Figure 2D). Moreover, we observed that plasma from PI donors better recognized HCoV-HKU1 S than plasma from naïve donors at every time point studied, suggesting that natural infection induced cross reactive Abs more efficiently than vaccination.

We then evaluated the capacity of the different plasma samples to bind S from another highly pathogenic human coronavirus (SARS-CoV-1). We observed that plasma from PI individuals had Abs able to recognize to some extent SARS-CoV-1 S (Figure 2E). This is likely related the close genetic relationship between SARS-CoV-2 and SARS-CoV-1 (Rabaan et al., 2020; Sarkar et al., 2021). As previously observed (Tauzin et al., 2021), both vaccine doses significantly increased the level of recognition of the SARS-CoV-1 Spike in the naïve group (Figure 2E). In the PI group, only the first dose significantly improved the recognition. We note that the long interval between doses brings SARS-CoV-2 naïve individuals to recognize the different variant Spikes and related HCoV to the same extent than previously-infected individuals shortly after the second dose (V3) but followed by a decline to significantly lower levels than PI individuals at V4.

### Functional activities of vaccine-elicited antibodies

We (Tauzin et al., 2021) and others (Collier et al., 2021; Goel et al., 2021b; Planas et al., 2021b; Sahin et al., 2020) reported that three weeks post first Pfizer/BioNTech dose, SARS-CoV-2 S specific Abs with weak neutralizing properties are elicited. Nevertheless, these Abs present robust Fc-mediated effector functions as measured by their capacity to mediate antibody-dependent cellular cytotoxicity (ADCC) (Tauzin et al., 2021). To obtain a better understanding of this functional property over time, we tested all plasma samples with our previously reported ADCC assay (Anand et al., 2021; Beaudoin-Bussieres et al., 2021; Tauzin et al., 2021; Ullah et al., 2021). As expected, and in agreement with the absence of SARS-CoV-2 S specific Abs at baseline, no ADCC activity was observed for the naïve group before vaccination (Figure 3A). Plasma from the PI group maintained some levels of ADCC activity before vaccination, in agreement with a longitudinal study following immune responses in convalescent donors (Anand et al., 2021). Three weeks after the first dose, ADCC activity was elicited in both groups, but was significantly higher in the PI group. A decline in ADCC responses was observed in both groups nine weeks after V1 (V2, i.e., 12 weeks post vaccination). The second dose strongly boosted ADCC activity in the naïve group but remained stable for the PI groups. In agreement with the recognition of different hCoV Spikes presented in Figure 2, the capacity of PI to mediate ADCC remained relatively stable at V4 but significantly declined for naïve individuals. We note that the levels of ADCC activity were significantly higher in the PI group at all timepoints (Figure 3A).

**Figure 3.**
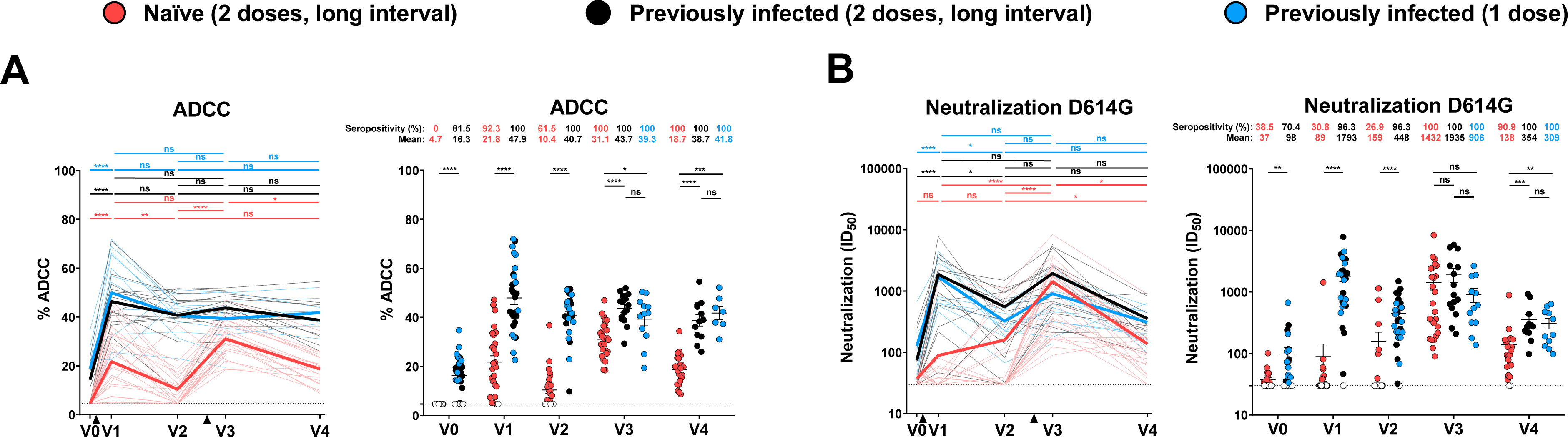
Fc-effector function and neutralization activities in SARS-CoV-2 naïve and previously-infected individuals before and after Pfizer/BioNTech mRNA vaccine. (**A**) CEM.NKr parental cells were mixed at a 1:1 ratio with CEM.NKr-Spike cells and were used as target cells. PBMCs from uninfected donors were used as effector cells in a FACS-based ADCC assay. (**B**) Neutralizing activity was measured by incubating pseudoviruses bearing SARS-CoV-2 S glycoproteins, with serial dilutions of plasma for 1 h at 37°C before infecting 293T-ACE2 cells. Neutralization half maximal inhibitory serum dilution (ID50) values were determined using a normalized non-linear regression using GraphPad Prism software. Naïve and PI donors with a long interval between the two doses are represented by red and black points respectively and PI donors who received just one dose by blue points. (**Left panels**) Each curve represents the values obtained with the plasma of one donor at every time point. Mean of each group is represented by a bold line. The time of vaccine dose injections is indicated by black triangles. (**Right panels**) Plasma samples were grouped in different time points (V0, V1, V2, V3 and V4). Undetectable measures are represented as white symbols, and limits of detection are plotted. Error bars indicate means ± SEM. (* P < 0.05; ** P < 0.01; *** P < 0.001; **** P < 0.0001; ns, non-significant).

Neutralizing activity in plasma is thought to play an important role in vaccine efficacy (Jackson et al., 2020; Muruato et al., 2020; Polack et al., 2020). Accordingly, it has been recently identified as an immune-correlate of protection in the mRNA-1273 COVID-19 vaccine efficacy trial (Gilbert et al., 2021). To evaluate the vaccine neutralizing response over time, we measured the capacity of plasma samples to neutralize pseudoviral particles carrying the SARS-CoV-2 S D614G glycoprotein (Figure 3B). We did not detect a significant increase in neutralization in plasma isolated three weeks post vaccination of the naïve group, as previously described (Tauzin et al., 2021). Interestingly, nine weeks later (V2, i.e., 12 weeks post vaccination), we observed increased neutralizing activity in a few donors (Figure 3B). All donors presented a significant increase in neutralizing activity three weeks after the second dose. Importantly, the level of neutralizing activity of double vaccinated naïve individuals reached the same levels than in the PI group after one or two doses. In this latter group (PI), we measured low neutralizing activity before vaccination, consistent with remaining neutralizing activity in convalescent donors after several months post symptoms onset (Anand et al., 2021; Gaebler et al., 2021; Tauzin et al., 2021). As previously described, the first dose strongly increased neutralization activity (Stamatatos et al., 2021; Tauzin et al., 2021), but this activity significantly decreased a few weeks after (V2, i.e., 12 weeks post vaccination). The second dose boosted the neutralizing activity to the levels reached three weeks after the first dose. No difference in neutralization was observed between V1 and V3 for PI individuals. In contrast, in naïve individuals we observed a significantly higher neutralizing activity after the second dose compared to the first one (Figure 3B). Thus, while one dose is required to reach maximum neutralization activity in PI individuals, this activity decays over time and a second dose is required to bring back its maximum potential. On the other hand, naïve individuals requires both doses to achieve the same level of PI vaccinated individuals three weeks after the second dose. However, the neutralizing activity declined more rapidly in the naïve group compared to PI individuals. Again, we observed no differences between PI that received one or two doses.

### Neutralizing activity against variants of concern

SARS-CoV-2 is evolving, and variants of concern are emerging globally (Davies et al., 2021; Prévost and Finzi, 2021; Sabino et al., 2021; Tegally et al., 2020; Volz et al., 2021). To evaluate whether the long interval between the two doses impacted the capacity of vaccine-elicited antibodies to neutralize VOCs and VOI, we measured the neutralizing activity against pseudoviral particles bearing selected variant Spikes (Figure S3). For all the variants tested, we observed a similar pattern than for the D614G S, with neutralizing Abs mainly induced after the second dose in the naïve group (Figure S3A-E). Previously-infected individuals followed a different pattern. While their plasma had some levels of neutralizing activity at baseline, it gained potency and breadth after the first dose. A second dose did not further enhance this activity.

We also noted that, with the exception of B.1.1.7, plasma from the PI group prior to vaccination (V0) neutralized less efficiently all pseudoviral particles bearing variant Spikes compared to the D614G (Figure S3A). Importantly, both doses boosted the neutralizing activity against all variants and SARS-CoV-1 Spike at V3 (Figure S2D). As observed with the D614G S, the neutralizing activity decreased at V4 for all VOCs tested (Figure S3E).

Vaccination of PI individuals was shown to increase neutralization against pseudoviral particles bearing the SARS-CoV-1 Spike (Stamatatos et al., 2021; Tauzin et al., 2021). This Spike is used as a representative variant that is even more dissimilar to the vaccine, which was based on the ancestral Wuhan strain. While only one dose was sufficient to provide SARS-CoV-1 neutralizing capacity in PI individuals, two were required in naïve individuals. Three weeks after the second dose (V3), plasma from naïve individuals reached the same level of neutralizing activity against pseudoviral particles bearing the SARS-CoV-1 Spike than PI. Thus, suggesting that the delayed boosting in naïve individuals allows antibody maturation resulting in enhanced breath (Figure S3).

### RBD avidity of vaccine-elicited antibodies

To gather evidence of vaccine-elicited antibodies maturation over time, we longitudinally followed RBD avidity. Briefly, we modified our ELISA assay by adding a chaotropic agent (8 M urea) to the washing buffer, as reported (Björkman et al., 1999; Fialová et al., 2017; Wang et al., 2019). By performing the ELISA assay in parallel, with washing steps having or not urea (see STAR Methods for details), we established an RBD avidity index (Figures 4 and S4), which provides an overall idea of the accumulated strength of vaccine-elicited antibodies affinities over time (Rudnick and Adams, 2009). For PI individuals, we observed that the first dose significantly increased the RBD avidity index. The second dose did not further improve the avidity. We observed no significant differences between PI donors who received one or two doses at V3 and V4. No RBD avidity index could be established at V0 for naïve individuals since they don’t have anti-RBD antibodies. However, the first dose elicited anti-RBD antibodies with a low RBD avidity index as compared to the PI group. Remarkably, the second dose increased RBD avidity to the same level than for PI individuals at V3 and remained relatively constant over time (V4, Figure 4).

**Figure 4.**
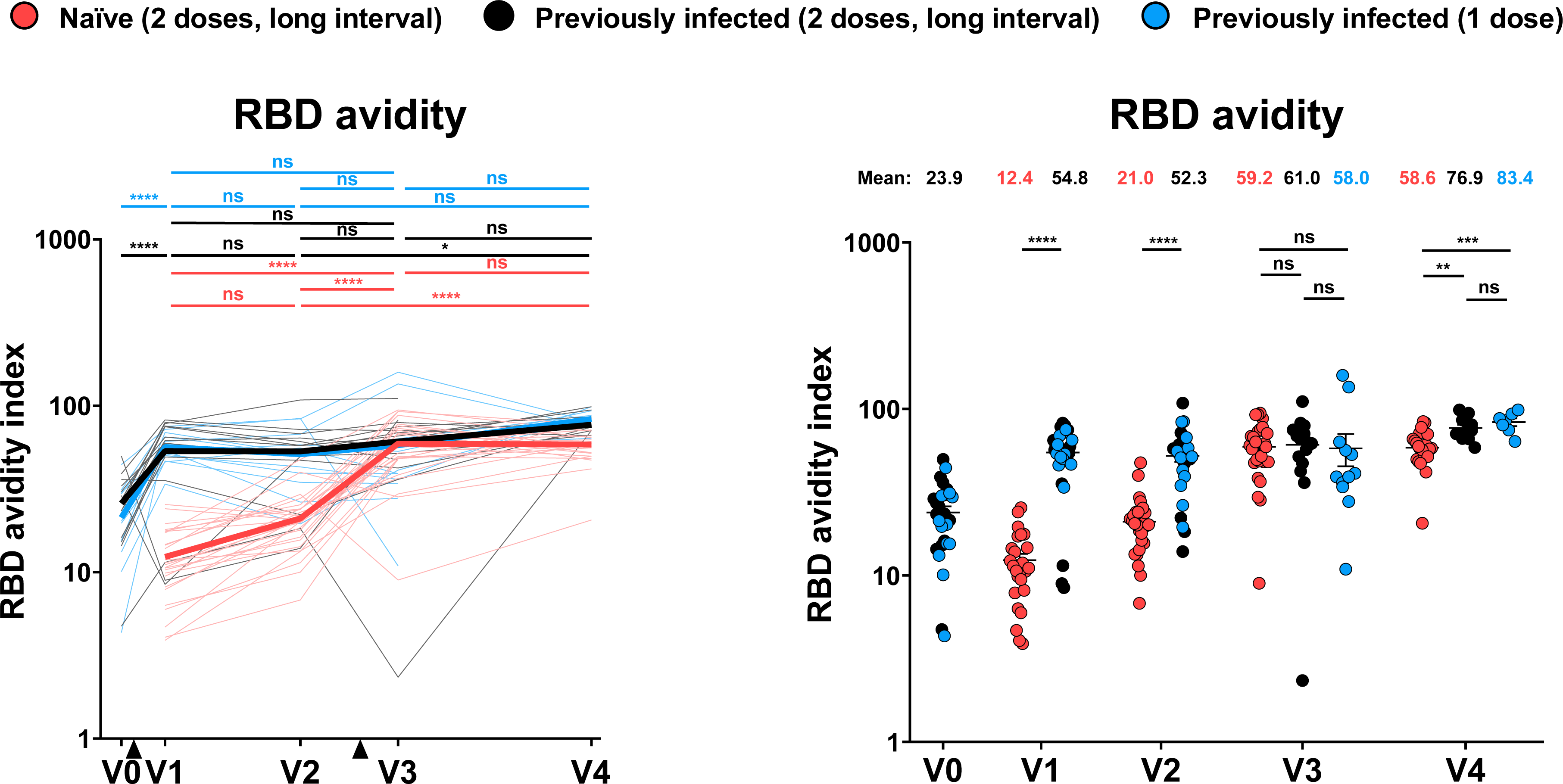
RBD avidity of vaccine-elicited antibodies. Indirect ELISA and stringent ELISA were performed by incubating plasma samples from naïve and PI donors collected at V0, V1, V2, V3 and V4 with recombinant SARS-CoV-2 RBD protein. Anti-RBD Ab binding was detected using HRP-conjugated anti-human IgG. Relative light unit (RLU) values obtained were normalized to the signal obtained with the anti-RBD CR3022 mAb present in each plate. The RBD avidity index corresponded to the value obtained with the stringent (8M urea) ELISA divided by that obtained with the ELISA. Naïve and PI donors with a long interval between the two doses are represented by red and black points respectively and PI donors who received just one dose by blue points. (**Left panels**) Each curve represents the values obtained with the plasma of one donor at every time point. Mean of each group is represented by a bold line. The time of vaccine dose injections is indicated by black triangles. (**Right panels**) Plasma samples were grouped in different time points (V0, V1, V2, V3 and V4). Undetectable measures are represented as white symbols, and limits of detection are plotted. Error bars indicate means ± SEM. (* P < 0.05; ** P < 0.01; *** P < 0.001; **** P < 0.0001; ns, non-significant).

### Humoral responses in individuals receiving a short dose interval regimen

We also analyzed the humoral responses of 12 SARS-CoV-2 naïve donors from a separate cohort who received their two doses of Pfizer/BioNTech mRNA vaccine four weeks apart (median [range]: 30 days [22–34 days]) (Table 1 and Figure 5A). For these donors, blood samples were only collected at V3, three weeks (median [range]: 24 days [12–37 days]) after the first dose, allowing a direct side-by-side comparison of humoral responses at V3 with our cohort of naïve individuals that received the two doses 16 weeks apart(Figure 1A and 5A). Naïve individuals that received the long interval regimen had more anti-RBD IgG (Figure 5B) and presented a significantly higher RBD-avidity index (Figure 5C) than naïve donors who received their two doses 4 weeks apart. We also observed major differences related to their capacity to recognize the full Spike of different variants. Plasma from short interval vaccinated individuals was significantly less efficient at recognizing the D614G S and all other S variants tested, except for the B.1.526 S (Figure 5D). Their capacity to mediate ADCC was also lower, albeit didn’t reach statistical significance (Figure 5E). Remarkably, the neutralization of pseudoviral particles bearing D614G or almost all the variant Spike tested was significantly lower for individuals that received the two doses with a short interval (Figure 5F). No neutralizing activity against SARS-CoV-1 was observed after a short interval (Figure 5F). In contrast, plasmas from naïve individual who received their two doses sixteen weeks apart presented a strong neutralizing activity against all the SARS-CoV-2 variants but also the SARS-CoV-1 pseudoviruses (Figure 5F).

**Figure 5.**
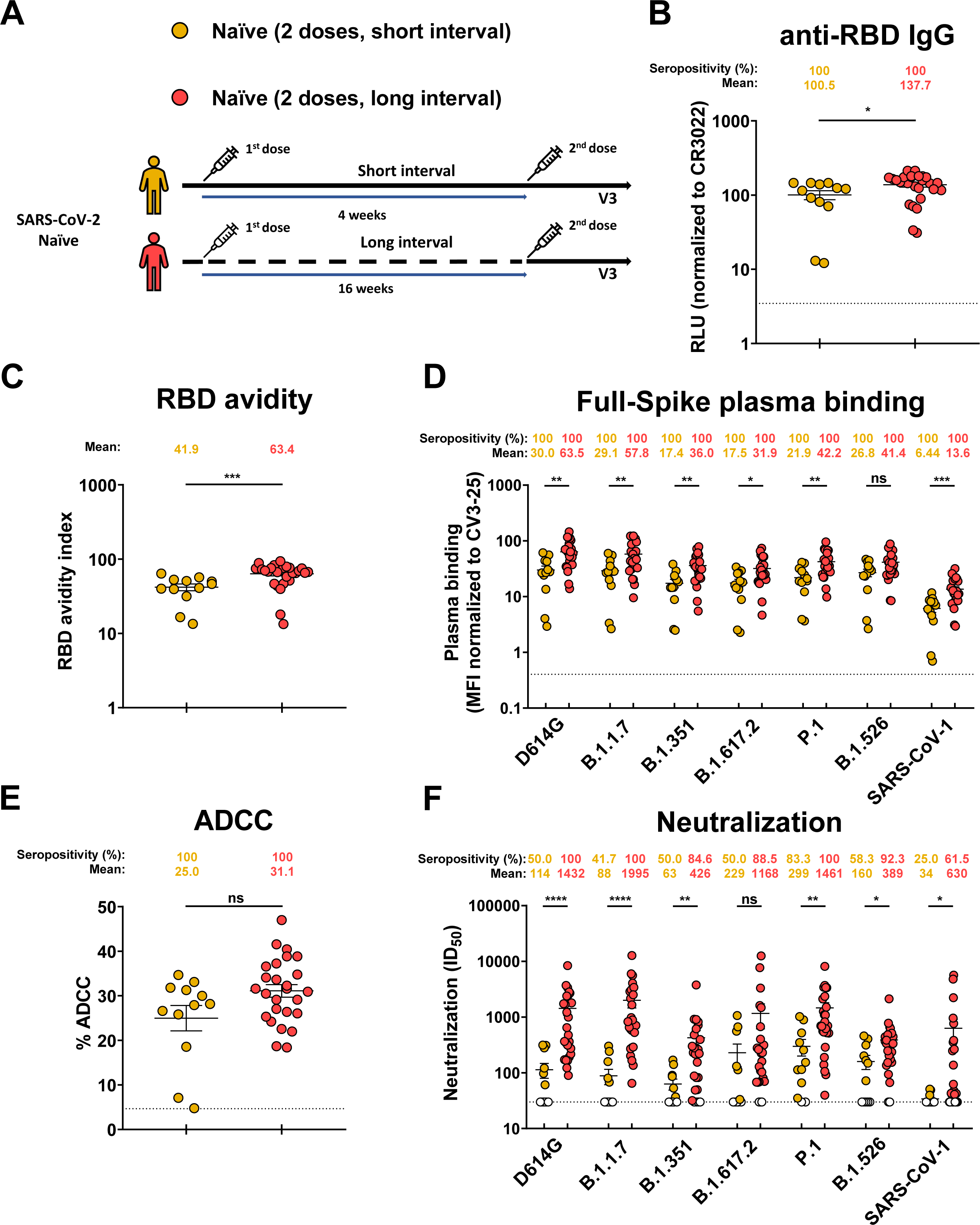
Humoral responses in SARS-CoV-2 naïve individuals that received a short dose versus a long dose interval. (**A**) SARS-CoV-2 vaccine cohort design. (**B**) Indirect ELISA was performed by incubating plasma samples from naïve donors collected at V3 with recombinant SARS-CoV-2 RBD protein. Anti-RBD Ab binding was detected using HRP-conjugated anti-human IgG. Relative light unit (RLU) values obtained with BSA (negative control) were subtracted and further normalized to the signal obtained with the anti-RBD CR3022 mAb present in each plate. (**C**) Indirect ELISA and stringent ELISA was performed by incubating plasma samples from naïve donors collected at V3 with recombinant SARS-CoV-2 RBD protein. Anti-RBD Ab binding was detected using HRP-conjugated anti-human IgG. Relative light unit (RLU) values obtained were normalized to the signal obtained with the anti-RBD CR3022 mAb present in each plate. RBD avidity index corresponded to the value obtained with the stringent ELISA divided by that obtained with the ELISA. (**D**) 293T cells were transfected with the indicated full-length S and stained with the CV3-25 Ab or with plasma from naïve donors collected at V3 and analyzed by flow cytometry. The values represent the MFI normalized by CV3-25 Ab binding. (**E**) CEM.NKr parental cells were mixed at a 1:1 ratio with CEM.NKr-Spike cells and were used as target cells. PBMCs from uninfected donors were used as effector cells in a FACS-based ADCC assay. (**F**) Neutralizing activity was measured by incubating pseudoviruses bearing SARS-CoV-2 S glycoproteins or SARS-CoV-1 S glycoprotein, with serial dilutions of plasma for 1 h at 37°C before infecting 293T-ACE2 cells. Neutralization half maximal inhibitory serum dilution (ID50) values were determined using a normalized non-linear regression using GraphPad Prism software. Naïve donors vaccinated with a short or a long interval between the two doses are represented by yellow or red points respectively. Plasma samples were grouped at V3. Undetectable measures are represented as white symbols, and limits of detection are plotted. Error bars indicate means ± SEM. (* P < 0.05; ** P < 0.01; *** P < 0.001; **** P < 0.0001; ns, non-significant).

### Integrated analysis of vaccine responses elicited with a sixteen-weeks interval between doses

When studying the network of pairwise correlations among all studied immune variables in SARS-CoV-2 naïve individuals (Figure 6A), we observed a sparsely interconnected network after the first vaccine dose with focused clusters among binding and neutralization responses, respectively. Over time, the network induced upon the 1^st^ vaccination slightly collapsed until the delayed 2^nd^ vaccination triggered a dense network of positive correlations involving binding, RBD avidity, neutralization responses against several SARS-CoV-2 variants and SARS-CoV-1, ADCC, and memory B cell responses. Importantly, this network remained associated 4 months after the second dose. As expected, for PIs individuals we observed an integrated network at baseline (i.e., before vaccination). Natural infection critically primes the quality of anti-SARS-CoV-2 humoral responses in infected hosts, and successive vaccination seems to increase certain titers (Figures 1-4) but does not essentially change the quality/relatedness of the induced responses. Associations remained relatively stable across all timepoints (Figure 6B).

**Figure 6.**
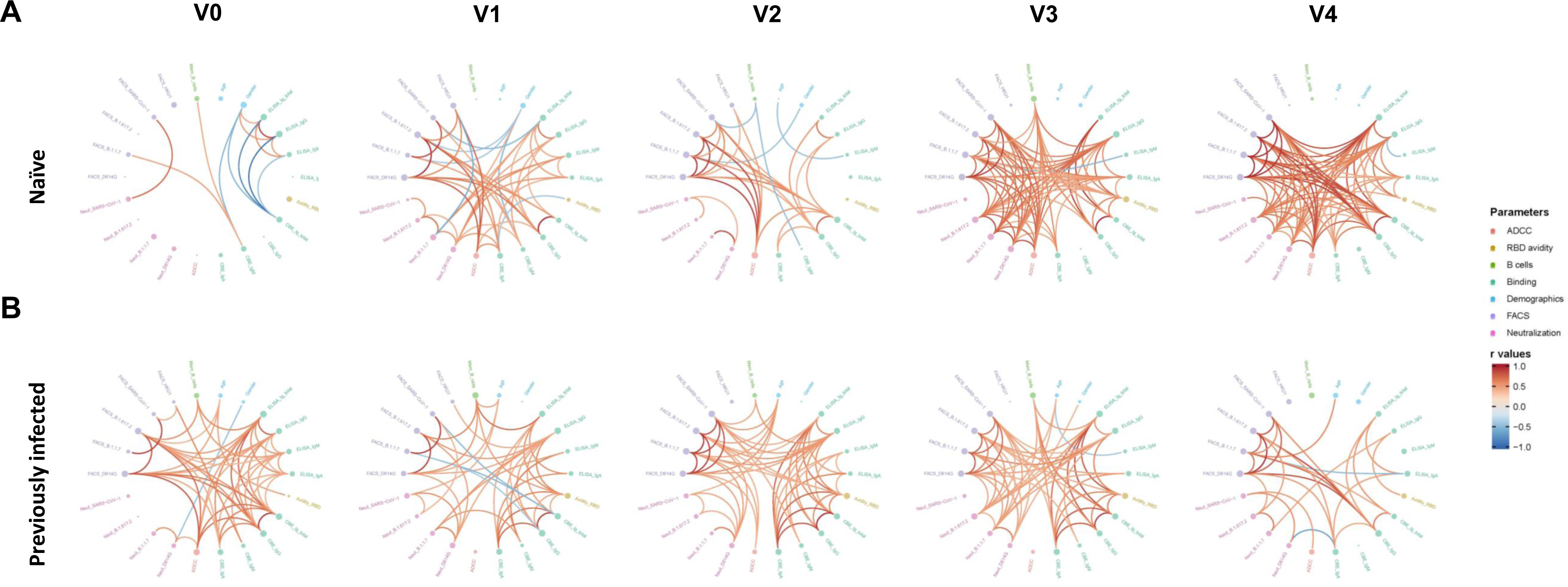
Mesh correlations of humoral response parameters in SARS-CoV-2 naïve and previously-infected individuals before and after Pfizer/BioNTech mRNA vaccine. Edge bundling correlation plots where red and blue edges represent positive and negative correlations between connected parameters, respectively. Only significant correlations (p < 0.05, Spearman rank test) are displayed. Nodes are color coded based on the grouping of parameters according to the legend. Node size corresponds to the degree of relatedness of correlations. Edge bundling plots are shown for correlation analyses using ten different datasets; i.e., SARS-CoV-2 naive (**A**) or previously infected (**B**) individuals at V0, V1, V2, V3 and V4 respectively.

## DISCUSSION

The approved regimen of the BNT162b2 mRNA vaccine is the administration of two doses within a short interval of 3-4 weeks. Despite the rapid approval of different vaccine platforms, generating the required doses to immunize the world population represents a daunting task (Moore and Klasse, 2020). Confronted to vaccine scarcity, some jurisdictions decided to increase the interval between doses in order to increase the number of immunized individuals. This decision led to concerns about vaccine efficacy, notably against emergent variants rapidly spreading worldwide and more resistant notably against neutralizing Abs induced by vaccination (Annavajhala et al., 2021; Planas et al., 2021a; Puranik et al., 2021; Wall et al., 2021; Wang et al., 2021a). Here, we measured the humoral responses of SARS-CoV-2 naïve and SARS-CoV-2 PI individuals who received their two doses sixteen weeks apart.

We observed that in the SARS-CoV-2 naïve group the BNT162b2 mRNA vaccine elicited antibodies with weak avidity for the RBD and neutralizing activity but strong Fc-mediated functions three weeks after the first dose (Tauzin et al., 2021). These functional responses declined in the following weeks in the absence of a boost. This is consistent with an overall decline in the anti-RBD and anti-Spike antibodies before the delayed boost. However, our results support antibody maturation during this same period with a significant increase in RBD avidity. Administration of the second dose sixteen weeks later strongly enhanced antibody levels but also functional responses, notably neutralization against some VOCs/VOIs and even the divergent SARS-CoV-1. Therefore, despite initial concerns, the long interval between the doses did not result in poor immune responses, A limitation of our study is the relatively low number of individuals analyzed however, we note that our results are in agreement with recent findings (Parry et al., 2021; Payne et al., 2021). Our results further support the conclusions of a recent study suggesting that extending the interval between first and second doses may have optimized booster dose protection in Canada (Skowronski et al., 2021). The idea behind the strategy of delaying the second dose was to provide some level of immunity to a larger number of individuals than if the second dose would have been saved to administer them three weeks later. However, despite the immunological benefits of increasing the interval between the two doses, this also increases the probability of being infected before the boost.

Several studies have shown that vaccination of previously-infected individuals elicits strong cellular and humoral responses (Efrati et al., 2021; Lozano-Ojalvo et al., 2021; Stamatatos et al., 2021; Tauzin et al., 2021; Urbanowicz et al., 2021). In agreement with these studies, we found that vaccination of these individuals resulted in the induction of strong humoral responses. These responses remained relatively stable over time. We noticed that the second dose did not result in a significant enhancement of these responses, even with a long interval of 16 weeks between doses. Our results demonstrate that, while the second dose boosts the humoral response, PI individuals reach their peak of immunity after the first dose and these responses remained relatively stable for at least 8 months. Altogether, these results suggest that a second dose for PI individuals might be delayed beyond sixteen weeks after the first dose. These observations are in agreement with recent studies showing that PI individuals had maximal humoral and CD4+ and CD8+ T cell responses after the first dose of an mRNA vaccine; the second did not strongly boost these responses (Goel et al., 2021a; Lozano-Ojalvo et al., 2021; Painter et al., 2021).

In contrast, here we show that a delayed second vaccine boost in naïve individuals significantly enhances several immune responses and tightens the network of linear correlations among those. The involved immune variables were humoral and cellular responses directed against SARS-CoV-2, including diverse variants, and SARS-CoV-1, but not or marginally against HCoV-HKU1. Thus, the potency, quality, and concerted triggering of immune responses appear enhanced in naïve individuals vaccinated with a prolonged interval of 16 weeks between first and second shot. Shortly after the boost, these responses were comparable to those obtained after vaccination of previously infected individuals. However, these responses declined more rapidly in naïve individuals than in PI individuals, suggesting the natural infection associated with vaccination leads to a longer immunity.

We also analyzed humoral responses in a cohort of naïve donors who received their two doses according to the approved short three-four-week interval. Plasma collected three weeks post second dose, had significantly lower humoral activities notably neutralizing activity against D614G strain and some VOCs/VOIs compared to naïve donors receiving the long interval. These results are in agreement with recent studies showing that increasing the interval between the two doses led to significant higher immune responses and vaccine effectiveness (Payne et al., 2021; Skowronski et al., 2021). Importantly, we observed a significant difference in the RBD avidity of the IgG, suggesting that increasing the time between the two doses facilitates antibody maturation, consistent with a better maturation of B cells in the germinal center (Kim et al., 2021).

Field effectiveness studies in Israël and the USA, where a short interval between doses is recommended, suggest waning protection of the BNT162b2 mRNA vaccine series against non-severe disease after a period of approximately 5 months (CDC, 2021; Goldberg et al., 2021; JCVI, 2021; Tartof et al., 2021). However, SARS-CoV-2 specific memory B cell and CD4+ T cell responses remains stable for the following 6 months, likely protecting from severe disease (Goel et al., 2021a). It will be of critical importance to monitor immune responses and vaccine effectiveness of extended vaccine schedules over time. If the strong humoral response seen with this extended schedule is longer-lasting than immune responses following the authorized schedule, the need of a third dose might be delayed and this could have significant implications regarding control of COVID-19.

To end this pandemic, it will be necessary to rapidly vaccinate the world’s population, including in countries where vaccines are poorly available. The research community around the globe rapidly generated a wealth of data related to vaccine-elicited immune responses and vaccine efficacy. Globally, these results suggest that the current vaccine strategy that was initially deployed could be improved. Our results suggest that modifying the interval at which the two doses are administered might be an important factor to take into account. It will be important to keep in mind that a fine balance needs to be achieved in order to avoid infection between the two doses and at the same time provide sufficient time to elicit optimal humoral responses.

## Data Availability

This article includes all datasets generated and analyzed for this study. Further information and requests for resources and reagents should be directed to and will be fulfilled by the Lead Contact Author.

## ACKNOWLEDGMENTS

The authors are grateful to the donors who participated in this study. The authors thank the CRCHUM BSL3 and Flow Cytometry Platforms for technical assistance. We thank Dr. Stefan Pöhlmann (Georg-August University, Germany) for the plasmid coding for SARS-CoV-2 and SARS-CoV-1 S glycoproteins and Dr. M. Gordon Joyce (U.S. MHRP) for the monoclonal antibody CR3022. We also thank Amélie Boivin and Yves Grégoire at Héma-Québec for helping to access the samples from the PLASCOV Biobank and all the plasma donors who participate in this biobank. Plasma Donor Biobank at Hema-Quebec was supported by funding from the Public Health Agency of Canada, through the Vaccine Surveillance Reference group and the COVID-19 Immunity Task Force. The views expressed here do not necessarily reflect those of the Public Health Agency of Canada. The graphical abstract was prepared using images from <u>BioRender.com</u>. This work was supported by le Ministère de l’Économie et de l’Innovation du Québec, Programme de soutien aux organismes de recherche et d’innovation to A.F. and by the Fondation du CHUM. This work was also supported by a CIHR foundation grant #352417, by a CIHR operating Pandemic and Health Emergencies Research grant #177958, a CIHR stream 1 and 2 for SARS-CoV-2 Variant Research to A.F., and by an Exceptional Fund COVID-19 from the Canada Foundation for Innovation (CFI) #41027 to A.F. and D.E.K. Work on variants presented was also supported by the Sentinelle COVID Quebec network led by the LSPQ in collaboration with Fonds de Recherche du Québec Santé (FRQS) to A.F. This work was also partially supported by a CIHR COVID-19 rapid response grant (OV3 170632) and CIHR stream 1 SARS-CoV-2 Variant Research to MC. A.F. is the recipient of Canada Research Chair on Retroviral Entry no. RCHS0235 950-232424. MC is a Tier II Canada Research Chair in Molecular Virology and Antiviral Therapeutics. V.M.L. is supported by a FRQS Junior 1 salary award. D.E.K. is a FRQS Merit Research Scholar. G.B.B. is the recipient of a FRQS PhD fellowship and J.P. is the recipient of a CIHR PhD fellowship. G.S. is supported by a scholarship from the Department of Microbiologie, Infectiologie et Immunology of Université de Montréal. R.G. and A.L. were supported by MITACS Accélération postdoctoral fellowships. The funders had no role in study design, data collection and analysis, decision to publish, or preparation of the manuscript. We declare no competing interests.

## AUTHOR CONTRIBUTIONS

A.T. and A.F. conceived the study. A.T., G.B.B., R.G., J.Prévost, M.N., J.R., D.E.K., and A.F. designed experimental approaches. A.T., S.Y.G., G.B.B., D.V., R.G.,L.N., L.M., M.Benlarbi, D.C., M.N., A.L., J.Prévost, M.Boutin, G.S., A.N., C.B., Y.B., M.D., D.E.K., and A.F. performed, analyzed, and interpreted the experiments. A.T. and R.D. performed statistical analysis. S.Y.G., G.B.B., A.L., J.Prévost, G.G.L., H.M., G.G., Y.B., J.R., M.C and A.F. contributed unique reagents. J.Perreault, L.G., C.M., P.A., R.B., R.R., G.C.M., C.T. and V.M.-L. collected and provided clinical samples. G.D.S., and N.B. provided scientific input related to VOC and vaccine efficacy. A.T., and A.F. wrote the manuscript with inputs from others. Every author has read, edited, and approved the final manuscript.

## STAR METHODS

### RESOURCE AVAILABILITY

#### Lead contact

Further information and requests for resources and reagents should be directed to and will be fulfilled by the lead contact, Andrés Finzi

#### Materials availability

All unique reagents generated during this study are available from the Lead contact without restriction.

#### Data and code availability

The published article includes all datasets generated and analyzed for this study. Further information and requests for resources and reagents should be directed to and will be fulfilled by the Lead Contact Author.

### EXPERIMENTAL MODEL AND SUBJECT DETAILS

#### Ethics Statement

All work was conducted in accordance with the Declaration of Helsinki in terms of informed consent and approval by an appropriate institutional board. Blood samples were obtained from donors who consented to participate in this research project at CHUM (19.381) and from plasma donors who consented to participate in the Plasma Donor Biobank at Hema-Quebec (PLASCOV; REB-B-6-002-2021-003). Plasma and PBMCs were isolated by centrifugation and Ficoll gradient, and samples stored at -80°C and in liquid nitrogen, respectively, until use.

#### Human subjects

No specific criteria such as number of patients (sample size), clinical or demographic were used for inclusion, beyond PCR confirmed SARS-CoV-2 infection in adults.

#### Plasma and antibodies

Plasma from SARS-CoV-2 naïve and PI donors were collected, heat-inactivated for 1 hour at 56°C and stored at -80°C until ready to use in subsequent experiments. Plasma from uninfected donors collected before the pandemic were used as negative controls and used to calculate the seropositivity threshold in our ELISA, cell-based ELISA, ADCC and flow cytometry assays (see below). The RBD-specific monoclonal antibody CR3022 was used as a positive control in our ELISA, cell-based ELISA, and flow cytometry assays and was previously described (Anand et al., 2020; Beaudoin-Bussières et al., 2020; Meulen et al., 2006; Prévost et al., 2020). Horseradish peroxidase (HRP)-conjugated Abs able to detect all Ig isotypes (anti-human IgM+IgG+IgA; Jackson ImmunoResearch Laboratories) or specific for the Fc region of human IgG (Invitrogen), the Fc region of human IgM (Jackson ImmunoResearch Laboratories) or the Fc region of human IgA (Jackson ImmunoResearch Laboratories) were used as secondary Abs to detect Ab binding in ELISA and cell-based ELISA experiments. Alexa Fluor-647-conjugated goat anti-human Abs able to detect all Ig isotypes (anti-human IgM+IgG+IgA; Jackson ImmunoResearch Laboratories) were used as secondary Ab to detect plasma binding in flow cytometry experiments.

#### Cell lines

293T human embryonic kidney and HOS cells (obtained from ATCC) were maintained at 37°C under 5% CO_2_ in Dulbecco’s modified Eagle’s medium (DMEM) (Wisent) containing 5% fetal bovine serum (FBS) (VWR) and 100 μg/ml of penicillin-streptomycin (Wisent). CEM.NKr CCR5+ cells (NIH AIDS reagent program) were maintained at 37°C under 5% CO_2_ in Roswell Park Memorial Institute (RPMI) 1640 medium (Gibco) containing 10% FBS and 100 μg/ml of penicillin-streptomycin. 293T-ACE2 cell line was previously reported (Prévost et al., 2020). HOS and CEM.NKr CCR5+ cells stably expressing the SARS-CoV-2 S glycoproteins were previously reported (Anand et al., 2021).

### METHOD DETAILS

#### Plasmids

The plasmids expressing the human coronavirus Spike glycoproteins of SARS-CoV-2, SARS-CoV-1 (Hoffmann et al., 2013, 2020), HCoV-OC43 (Prévost et al., 2020) and MERS-CoV (Park et al., 2016) were previously reported. The HCoV-HKU1 S expressing plasmid was purchased from Sino Biological. The plasmids encoding the different SARS-CoV-2 Spike variants (D614G, B.1.1.7, B.1.351, P.1, B.1.526 and B.1.617.2) were previously described (Beaudoin-Bussières et al., 2020; Gong et al., 2021; Li et al., 2021; Tauzin et al., 2021).

#### Protein expression and purification

FreeStyle 293F cells (Invitrogen) were grown in FreeStyle 293F medium (Invitrogen) to a density of 1 x 10^6^ cells/mL at 37°C with 8 % CO_2_ with regular agitation (150 rpm). Cells were transfected with a plasmid coding for SARS-CoV-2 S RBD (Beaudoin-Bussières et al., 2020) using ExpiFectamine 293 transfection reagent, as directed by the manufacturer (Invitrogen). One week later, cells were pelleted and discarded. Supernatants were filtered using a 0.22 µm filter (Thermo Fisher Scientific). The recombinant RBD proteins were purified by nickel affinity columns, as directed by the manufacturer (Invitrogen). The RBD preparations were dialyzed against phosphate-buffered saline (PBS) and stored in aliquots at -80°C until further use. To assess purity, recombinant proteins were loaded on SDS-PAGE gels and stained with Coomassie Blue.

#### Enzyme-Linked Immunosorbent Assay (ELISA) and RBD avidity index

The SARS-CoV-2 RBD ELISA assay used was previously described (Beaudoin-Bussières et al., 2020; Prévost et al., 2020). Briefly, recombinant SARS-CoV-2 S RBD proteins (2.5 μg/ml), or bovine serum albumin (BSA) (2.5 μg/ml) as a negative control, were prepared in PBS and were adsorbed to plates (MaxiSorp Nunc) overnight at 4°C. Coated wells were subsequently blocked with blocking buffer (Tris-buffered saline [TBS] containing 0.1% Tween20 and 2% BSA) for 1h at room temperature. Wells were then washed four times with washing buffer (Tris-buffered saline [TBS] containing 0.1% Tween20). CR3022 mAb (50 ng/ml) or a 1/250 dilution of plasma were prepared in a diluted solution of blocking buffer (0.1 % BSA) and incubated with the RBD-coated wells for 90 minutes at room temperature. Plates were washed four times with washing buffer followed by incubation with secondary Abs (diluted in a diluted solution of blocking buffer (0.4% BSA)) for 1h at room temperature, followed by four washes. To calculate the RBD-avidity index, we performed a stringent ELISA, where the plates were washed with a chaotropic agent, 8M of urea, added of the washing buffer. HRP enzyme activity was determined after the addition of a 1:1 mix of Western Lightning oxidizing and luminol reagents (Perkin Elmer Life Sciences). Light emission was measured with a LB942 TriStar luminometer (Berthold Technologies). Signal obtained with BSA was subtracted for each plasma and was then normalized to the signal obtained with CR3022 present in each plate. The seropositivity threshold was established using the following formula: mean of pre-pandemic SARS-CoV-2 negative plasma + (3 standard deviation of the mean of pre-pandemic SARS-CoV-2 negative plasma).

#### Cell-Based ELISA

Detection of the trimeric SARS-CoV-2 S at the surface of HOS cells was performed by a previously-described cell-based enzyme-linked immunosorbent assay (ELISA) (Anand et al., 2021). Briefly, parental HOS cells or HOS-Spike cells were seeded in 96-well plates (4×10^4^ cells per well) overnight. Cells were blocked with blocking buffer (10 mg/ml nonfat dry milk, 1.8 mM CaCl2, 1 mM MgCl2, 25 mM Tris [pH 7.5], and 140 mM NaCl) for 30 min. CR3022 mAb (1 μg/ml) or plasma (at a dilution of 1/250) were prepared in blocking buffer and incubated with the cells for 1h at room temperature. Respective HRP-conjugated Abs were then incubated with the samples for 45 min at room temperature. For all conditions, cells were washed 6 times with blocking buffer and 6 times with washing buffer (1.8 mM CaCl2, 1 mM MgCl2, 25 mM Tris [pH 7.5], and 140 mM NaCl). HRP enzyme activity was determined after the addition of a 1:1 mix of Western Lightning oxidizing and luminol reagents (PerkinElmer Life Sciences). Light emission was measured with an LB942 TriStar luminometer (Berthold Technologies). Signal obtained with parental HOS was subtracted for each plasma and was then normalized to the signal obtained with CR3022 mAb present in each plate. The seropositivity threshold was established using the following formula: mean of pre-pandemic SARS-CoV-2 negative plasma + (3 standard deviation of the mean of pre-pandemic SARS-CoV-2 negative plasma).

#### Cell surface staining and flow cytometry analysis

293T cells were co-transfected with a GFP expressor (pIRES2-GFP, Clontech) in combination with plasmids encoding the full-length Spikes of SARS-CoV-2 variants or Spikes from different *Betacoronaviruses*. 48h post-transfection, S-expressing cells were stained with the CV3-25 Ab (Jennewein et al., 2021) or plasma (1/250 dilution). AlexaFluor-647-conjugated goat anti-human IgM+IgG+IgA Abs (1/800 dilution) were used as secondary Abs. The percentage of transfected cells (GFP+ cells) was determined by gating the living cell population based on viability dye staining (Aqua Vivid, Invitrogen). Samples were acquired on a LSRII cytometer (BD Biosciences) and data analysis was performed using FlowJo v10.7.1 (Tree Star). The seropositivity threshold was established using the following formula: (mean of pre-pandemic SARS-CoV-2 negative plasma + (3 standard deviation of the mean of pre-pandemic SARS-CoV-2 negative plasma). The conformational-independent S2-targeting mAb CV3-25 was used to normalize Spike expression. CV3-25 was shown to effectively recognize all SARS-CoV-2 Spike variants (Li et al., 2021).

#### ADCC assay

This assay was previously described (Anand et al., 2021). For evaluation of anti-SARS-CoV-2 antibody-dependent cellular cytotoxicity (ADCC), parental CEM.NKr CCR5+ cells were mixed at a 1:1 ratio with CEM.NKr cells stably expressing a GFP-tagged full length SARS-CoV-2 Spike (CEM.NKr.SARS-CoV-2.Spike cells). These cells were stained for viability (AquaVivid; Thermo Fisher Scientific, Waltham, MA, USA) and cellular dyes (cell proliferation dye eFluor670; Thermo Fisher Scientific) to be used as target cells. Overnight rested PBMCs were stained with another cellular marker (cell proliferation dye eFluor450; Thermo Fisher Scientific) and used as effector cells. Stained target and effector cells were mixed at a ratio of 1:10 in 96-well V-bottom plates. Plasma (1/500 dilution) or monoclonal antibody CR3022 (1 µg/mL) were added to the appropriate wells. The plates were subsequently centrifuged for 1 min at 300g, and incubated at 37°C, 5% CO_2_ for 5 hours before being fixed in a 2% PBS-formaldehyde solution. ADCC activity was calculated using the formula: [(% of GFP+ cells in Targets plus Effectors) - (% of GFP+ cells in Targets plus Effectors plus plasma/antibody)]/(% of GFP+ cells in Targets) x 100 by gating on transduced live target cells. All samples were acquired on an LSRII cytometer (BD Biosciences) and data analysis was performed using FlowJo v10.7.1 (Tree Star). The specificity threshold was established using the following formula: (mean of pre-pandemic SARS-CoV-2 negative plasma + (3 standard deviation of the mean of pre-pandemic SARS-CoV-2 negative plasma).

#### Virus neutralization assay

To produce the pseudoviruses, 293T cells were transfected with the lentiviral vector pNL4.3 R-E-Luc (NIH AIDS Reagent Program) and a plasmid encoding for the indicated S glycoprotein (D614G, B.1.1.7, P.1, B.1.351, B.1.617.2, B.1.526 and SARS-CoV) at a ratio of 10:1. Two days post-transfection, cell supernatants were harvested and stored at -80°C until use. For the neutralization assay, 293T-ACE2 target cells were seeded at a density of 1×10^4^ cells/well in 96-well luminometer-compatible tissue culture plates (Perkin Elmer) 24h before infection.

Pseudoviral particles were incubated with several plasma dilutions (1/50; 1/250; 1/1250; 1/6250; 1/31250) for 1h at 37°C and were then added to the target cells followed by incubation for 48h at 37°C. Then, cells were lysed by the addition of 30 µL of passive lysis buffer (Promega) followed by one freeze-thaw cycle. An LB942 TriStar luminometer (Berthold Technologies) was used to measure the luciferase activity of each well after the addition of 100 µL of luciferin buffer (15mM MgSO4, 15mM KPO4 [pH 7.8], 1mM ATP, and 1mM dithiothreitol) and 50 µL of 1mM d-luciferin potassium salt (Prolume). The neutralization half-maximal inhibitory dilution (ID50) represents the plasma dilution to inhibit 50% of the infection of 293T-ACE2 cells by pseudoviruses.

#### SARS-CoV-2-specific B cells characterization

To detect SARS-CoV-2-specific B cells, we conjugated recombinant RBD proteins with Alexa Fluor 488 or Alexa Fluor 594 (Thermo Fisher Scientific) according to the manufacturer’s protocol. Approximately 2×10^6^ frozen PBMCs from SARS-CoV-2 naïve and prior infection donors were prepared in Falcon® 5ml-round bottom polystyrene tubes at a final concentration of 4×10^6^ cells/mL in RPMI 1640 medium (GIBCO) supplemented with 10% of fetal bovine serum (Seradigm), Penicillin-Streptomycin (GIBCO) and HEPES (GIBCO). After a rest of 2h at 37°C and 5% CO_2_, cells were stained using Aquavivid viability marker (Biosciences) in DPBS (GIBCO) at 4°C for 20 min. The detection of SARS-CoV-2-antigen specific B cells was done by adding the RBD probes to the antibody cocktail (IgM BUV737, CD24 BUV805, IgG BV421, CD3 BV480, CD56 BV480, CD14 BV480, CD16 BV480, CD20 BV711, CD21 BV786, HLA DR BB700, CD27 APC R700; CD19 BV650, CD38 BB790, CD138 BUV661, CCR10 BUV395, IgD BUV563 and IgA PE). Staining was performed at 4°C for 30 min and cells were fixed using 1% paraformaldehyde (Sigma-Aldrich) at 4°C for 15 min. Stained PBMC samples were acquired on FACSymphony™ A5 Cell Analyzer (BD Biosciences) and analyzed using FlowJo v10.7.1 software.

### QUANTIFICATION AND STATISTICAL ANALYSIS

#### Statistical analysis

Symbols represent biologically independent samples from SARS-CoV-2 naïve individuals or SARS-CoV-2 PI individuals. Lines connect data from the same donor. Statistics were analyzed using GraphPad Prism version 8.0.1 (GraphPad, San Diego, CA). Every dataset was tested for statistical normality and this information was used to apply the appropriate (parametric or nonparametric) statistical test. Differences in responses for the same patient before and after vaccination were performed using Kruskal-Wallis tests. Differences in responses between naïve and PI individuals at each time point were measured by Mann-Whitney (V0, V1 and V2) or Kruskal-Wallis (V3 and V4) tests. Differences in responses against the different Spikes for the same patient were measured by Friedman tests. P values < 0.05 were considered significant; significance values are indicated as ∗ p < 0.05, ∗∗ p < 0.01, ∗∗∗ p < 0.001, ∗∗∗∗ p < 0.0001. Spearman’s R correlation coefficient was applied for correlations. Statistical tests were two-sided and p < 0.05 was considered significant.

#### Software scripts and visualization

Edge bundling graphs were generated in undirected mode in R and RStudio using ggraph, igraph, tidyverse,and RColorBrewer packages (R; R studio). Edges are only shown if p < 0.05, and nodes are sized according to the connecting edges’ r values. Nodes are color-coded according to groups of parameters.

## SUPPLEMENTAL INFORMATION

Supplemental information can be found online at …

---

**Figure S1:**
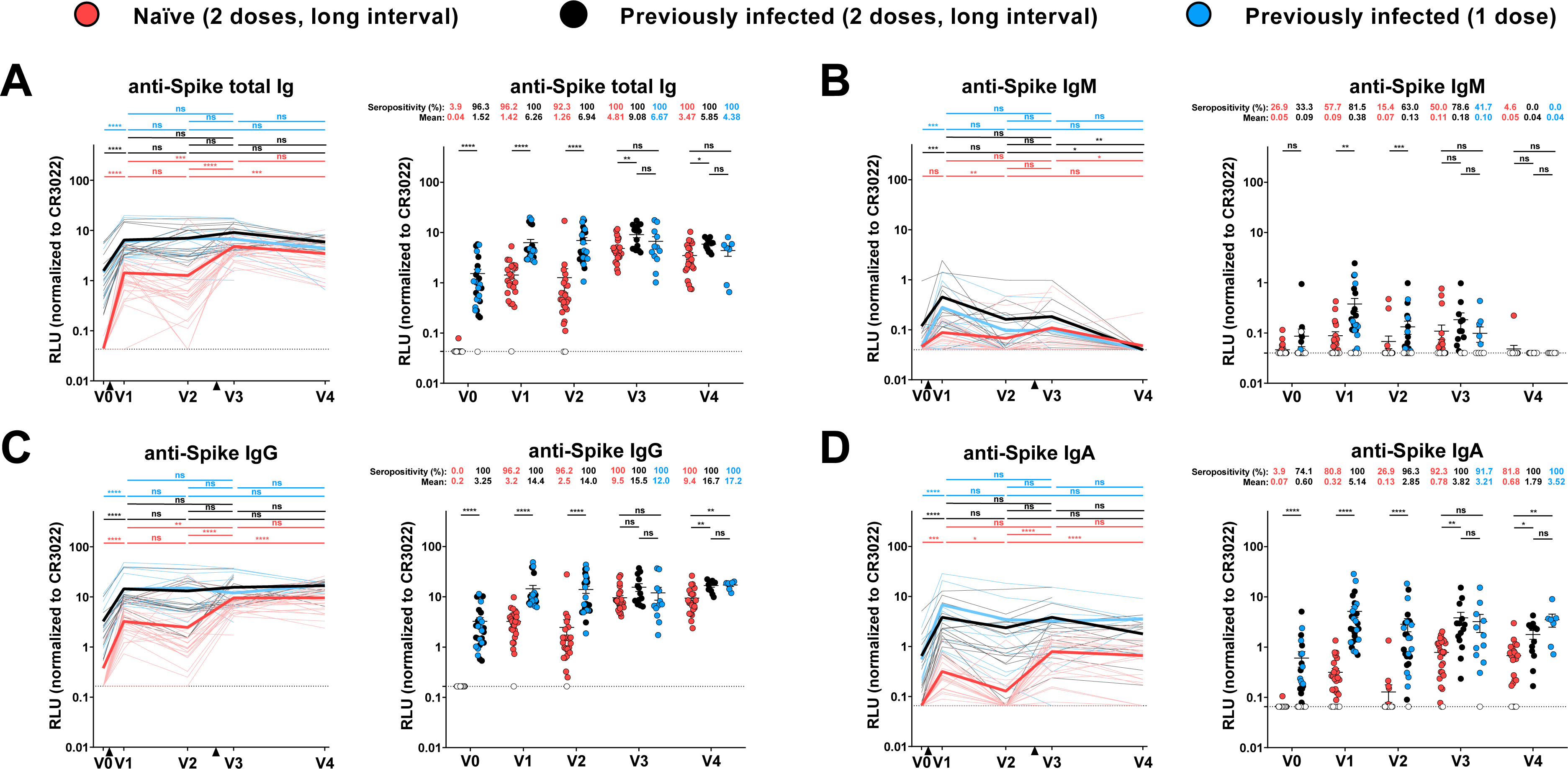
Elicitation of Spike-specific antibodies in SARS-CoV-2 naïve and previously-infected individuals, Related to Figures 1 and 6. (**A-D**) Cell-based ELISA was performed by incubating plasma samples from naïve and PI donors collected at V0, V1, V2, V3 and V4 with HOS cells expressing full-length SARS-CoV-2 S. Anti-S Ab binding was detected using HRP-conjugated (**A**) anti-human IgM+IgG+IgA (**B**) anti-human IgM, (**C**) anti-human IgG, or (**D**) anti-human IgA. RLU values obtained with parental HOS (negative control) were subtracted and further normalized to the signal obtained with the CR3022 mAb present in each plate. Naïve and PI donors with a long interval between the two doses are represented by red and black points respectively and PI donors who received just one dose by blue points. (Left panels) Each curve represents the normalized RLUs obtained with the plasma of one donor at every time point. Mean of each group is represented by a bold line. The time of vaccine dose injections is indicated by black triangles. (Right panels) Plasma samples were grouped in different time points (V0, V1, V2, V3 and V4). Undetectable measures are represented as white symbols, and limits of detection are plotted. Error bars indicate means ± SEM. (* P < 0.05; ** P < 0.01; *** P < 0.001; **** P < 0.0001; ns, non-significant).

**Figure S2:**
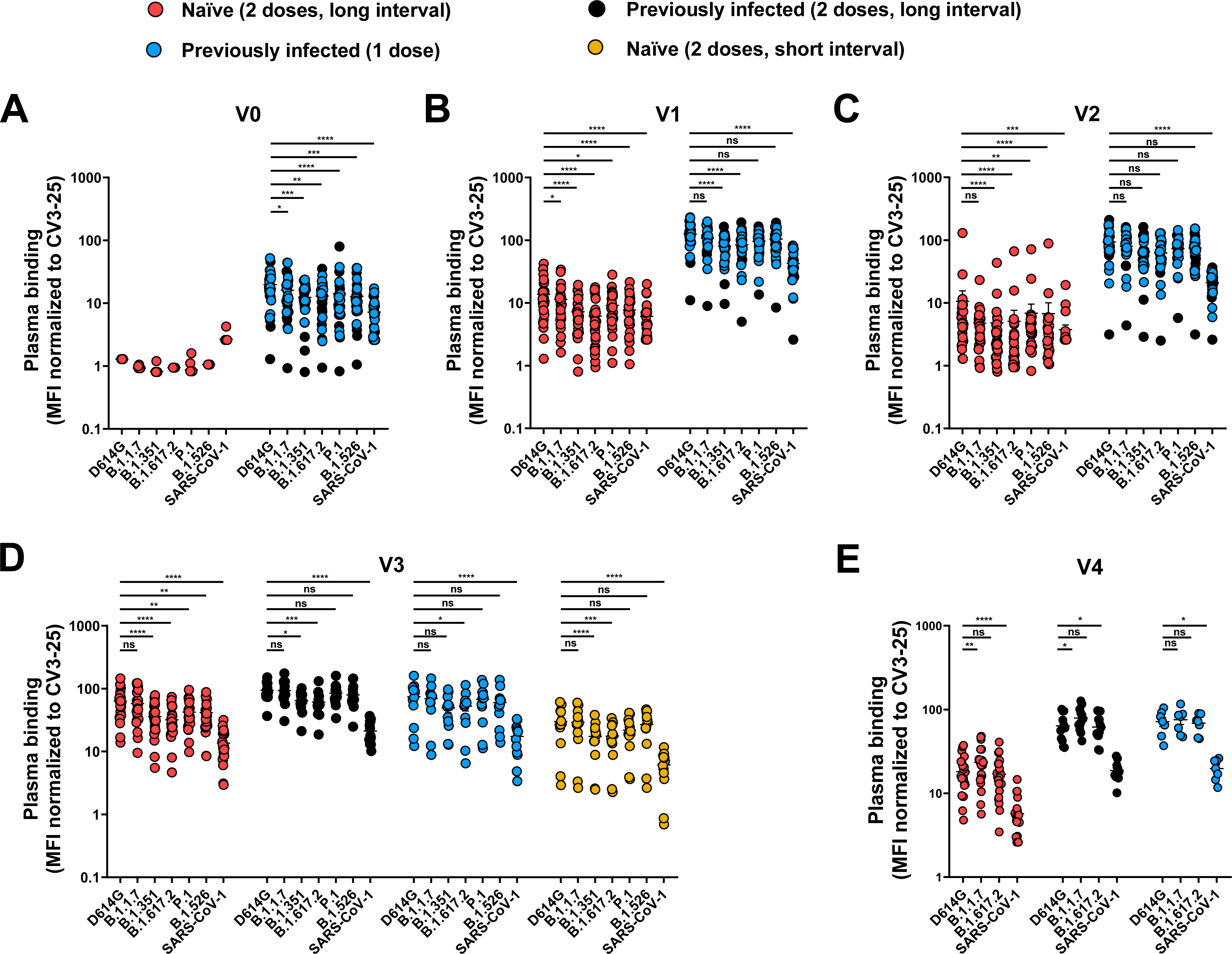
Recognition of SARS-CoV-2 Spike variants and SARS-CoV-1 Spike by plasma from naïve and PI donors at each time point, Related to Figures 2, 5 and 6. 293T cells were transfected with the indicated *Betacoronavirus* Spike and stained with the CV3-25 Ab or with plasma collected at V0 (**A**), V1 (**B**), V2 (**C**), V3 (**D**) and V4 (**E**) and analyzed by flow cytometry. Plasma recognitions are normalized with CV3-25 binding. Naïve and PI donors with a long interval between the two doses are represented by red and black points respectively, PI donors who received just one dose by blue points and naïve donors with a short interval between the two doses by yellow points. Error bars indicate means ± SEM. (* P < 0.05; ** P < 0.01; *** P < 0.001; **** P < 0.0001; ns, non-significant).

**Figure S3:**
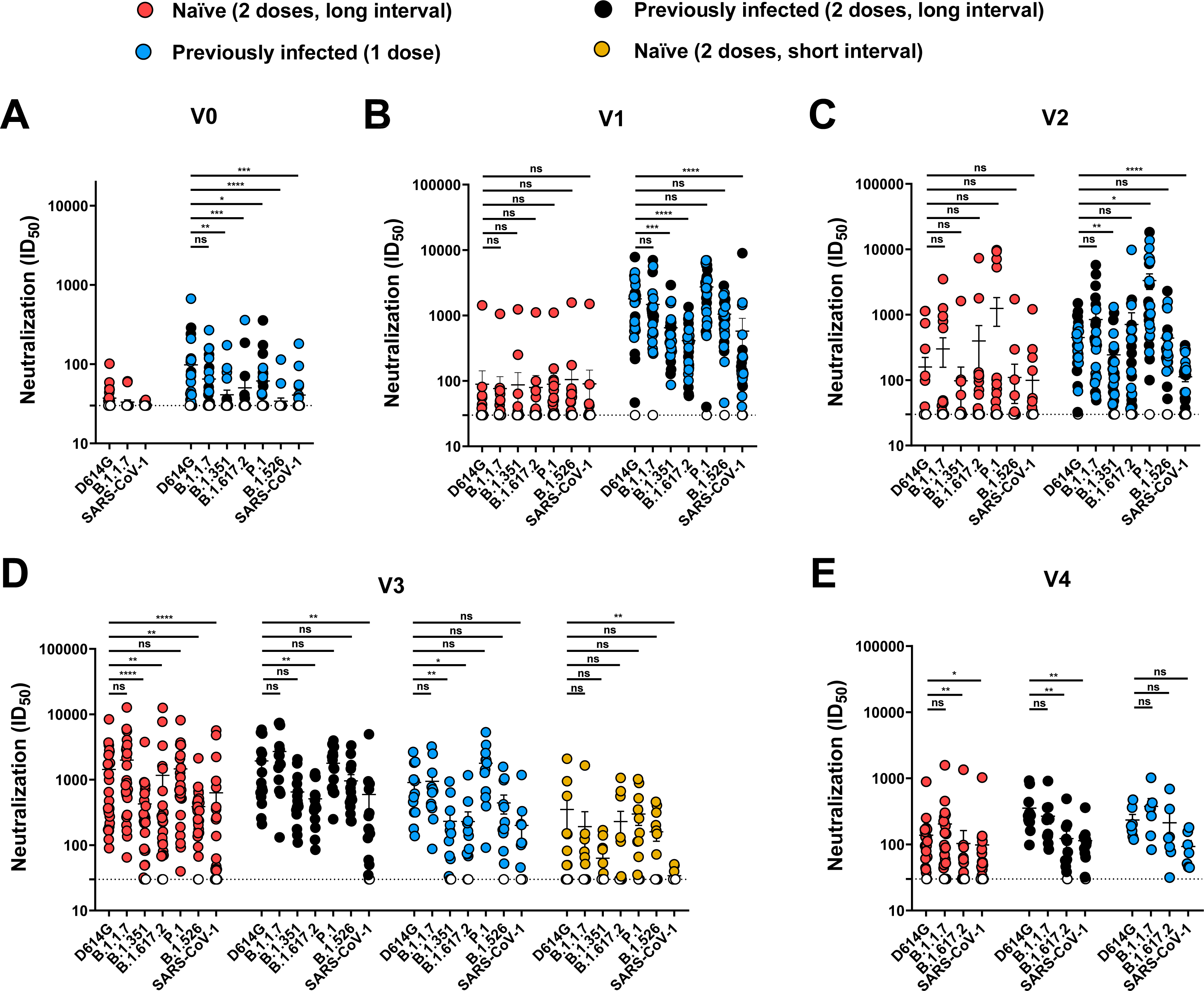
Neutralization of SARS-CoV-2 Spike variants and SARS-CoV-1 Spike by plasma from naïve and PI donors at each time point, Related to Figures 3, 5 and 6. Neutralizing activity was measured by incubating pseudoviruses bearing SARS-CoV-2 S variant or SARS-CoV-1 S glycoproteins, with serial dilutions of plasma collected at V0 (**A**), V1 ( **B**), V2 ( **C**), V3 ( **D**) and V4 (**E**) for 1 h at 37°C before infecting 293T-ACE2 cells. Neutralization half maximal inhibitory serum dilution (ID50) values were determined using a normalized non-linear regression using GraphPad Prism software. Naïve and PI donors with a long interval between the two doses are represented by red and black points respectively, PI donors who received just one dose by blue points and naïve donors with a short interval between the two doses by yellow points. Undetectable measures are represented as white symbols, and limits of detection are plotted. Error bars indicate means ± SEM. (* P < 0.05; ** P < 0.01; *** P < 0.001; **** P < 0.0001; ns, non-significant).

**Figure S4:**
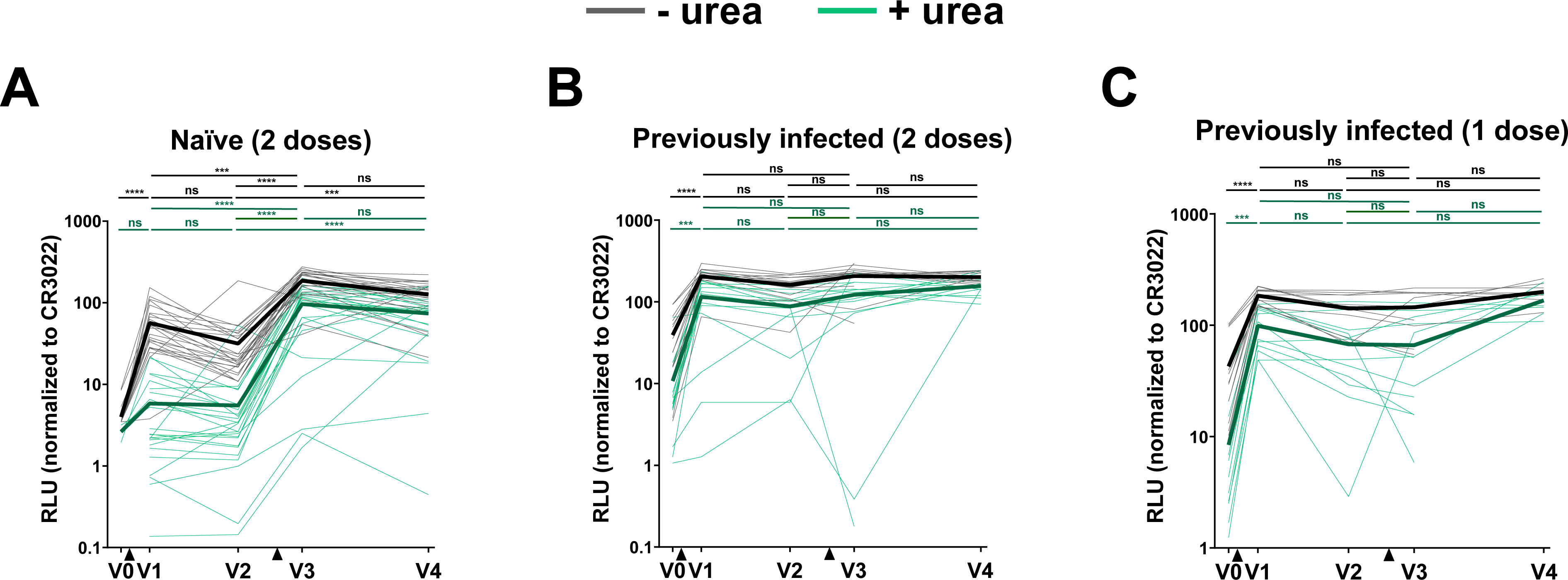
Comparison of the detection of RBD specific antibodies between ELISA and stringent ELISA in SARS-CoV-2 naïve and previously infected individuals, Related to Figure 4. (**A-C**) Indirect ELISA was performed by incubating plasma samples from naïve (**A**) PI vaccinated with two (**B**) or one dose (**C**) donors collected at V0, V1, V2, V3 and V4 with recombinant SARS-CoV-2 RBD protein. Anti-RBD Ab binding was detected using HRP-conjugated anti-human IgG. Relative light unit (RLU) values obtained were normalized to the signal obtained with the anti-RBD CR3022 mAb present in each plate. For ELISA (black curves), all the wash steps were made with washing buffer and for stringent ELISA (green curves), the wash steps were made with 8M of urea. Each curve represents the normalized RLUs obtained with the plasma of one donor at every time point. Mean of each group is represented by a bold line. The time of vaccine dose injections is indicated by black triangles. Error bars indicate means ± SEM. (* P < 0.05; ** P < 0.01; *** P < 0.001; **** P < 0.0001; ns, non-significant).

## REFERENCES

Allen, H., Vusirikala, A., Flannagan, J., Twohig, K.A., Zaidi, A., Chudasama, D., Lamagni, T., Groves, N., Turner, C., Rawlinson, C., et al. (2021). Household transmission of COVID-19 cases associated with SARS-CoV-2 delta variant (B.1.617.2): national case-control study. Lancet Reg. Health – Eur. 0.

Anand, S.P., Prévost, J., Richard, J., Perreault, J., Tremblay, T., Drouin, M., Fournier, M.-J., Lewin, A., Bazin, R., and Finzi, A. (2020). High-throughput detection of antibodies targeting the SARS-CoV-2 Spike in longitudinal convalescent plasma samples (Microbiology).

Anand, S.P., Prévost, J., Nayrac, M., Beaudoin-Bussières, G., Benlarbi, M., Gasser, R., Brassard, N., Laumaea, A., Gong, S.Y., Bourassa, C., et al. (2021). Longitudinal analysis of humoral immunity against SARS-CoV-2 Spike in convalescent individuals up to eight months post-symptom onset. Cell Rep. Med. 100290.

Annavajhala, M.K., Mohri, H., Wang, P., Nair, M., Zucker, J.E., Sheng, Z., Gomez-Simmonds, A., Kelley, A.L., Tagliavia, M., Huang, Y., et al. (2021). Emergence and expansion of SARS-CoV-2 B.1.526 after identification in New York. Nature 597, 703–708.

Baden, L.R., El Sahly, H.M., Essink, B., Kotloff, K., Frey, S., Novak, R., Diemert, D., Spector, S.A., Rouphael, N., Creech, C.B., et al. (2021). Efficacy and Safety of the mRNA-1273 SARS-CoV-2 Vaccine. N. Engl. J. Med. 384, 403–416.

Beaudoin-Bussières, G., Laumaea, A., Anand, S.P., Prévost, J., Gasser, R., Goyette, G., Medjahed, H., Perreault, J., Tremblay, T., Lewin, A., et al. (2020). Decline of Humoral Responses against SARS-CoV-2 Spike in Convalescent Individuals. MBio 11.

Beaudoin-Bussieres, G., Chen, Y., Ullah, I., Prevost, J., Tolbert, W.D., Symmes, K., Ding, S., Benlarbi, M., Gong, S.Y., Tauzin, A., et al. (2021). An anti-SARS-CoV-2 non-neutralizing antibody with Fc-effector function defines a new NTD epitope and delays neuroinvasion and death in K18-hACE2 mice (Microbiology).

Björkman, C., Näslund, K., Stenlund, S., Maley, S.W., Buxton, D., and Uggla, A. (1999). An IgG Avidity ELISA to Discriminate between Recent and Chronic Neospora Caninum Infection. J. Vet. Diagn. Invest. 11, 41–44.

Brown, K.A., Tibebu, S., Daneman, N., Schwartz, K., Whelan, M., and Buchan, S. (2021). Comparative Household Secondary Attack Rates associated with B.1.1.7, B.1.351, and P.1 SARS-CoV-2 Variants. medRxiv 2021.06.03.21258302.

CDC (2021). Interim Clinical Considerations for Use of COVID-19 Vaccines | CDC, https://www.cdc.gov/vaccines/covid-19/clinical-considerations/covid-19-vaccines-us.html.

Collier, D.A., De Marco, A., Ferreira, I.A.T.M., Meng, B., Datir, R.P., Walls, A.C., Kemp, S.A., Bassi, J., Pinto, D., Silacci-Fregni, C., et al. (2021). Sensitivity of SARS-CoV-2 B.1.1.7 to mRNA vaccine-elicited antibodies. Nature 1–10.

Dagpunar, J. (2021). Interim estimates of increased transmissibility, growth rate, and reproduction number of the Covid-19 B.1.617.2 variant of concern in the United Kingdom. medRxiv 2021.06.03.21258293.

Dan, J.M., Mateus, J., Kato, Y., Hastie, K.M., Yu, E.D., Faliti, C.E., Grifoni, A., Ramirez, S.I., Haupt, S., Frazier, A., et al. (2021). Immunological memory to SARS-CoV-2 assessed for up to 8 months after infection. Science eabf4063.

Davies, N.G., Abbott, S., Barnard, R.C., Jarvis, C.I., Kucharski, A.J., Munday, J.D., Pearson, C.A.B., Russell, T.W., Tully, D.C., Washburne, A.D., et al. (2021). Estimated transmissibility and impact of SARS-CoV-2 lineage B.1.1.7 in England. Science 372.

Dong, E., Du, H., and Gardner, L. (2020). An interactive web-based dashboard to track COVID-19 in real time. Lancet Infect. Dis. 20, 533–534.

ECDC (2021). Risk of spread of new SARS-CoV-2 variants of concern in the EU/EEA - first update, https://www.ecdc.europa.eu/sites/default/files/documents/COVID-19-risk-related-to-spread-of-new-SARS-CoV-2-variants-EU-EEA-first-update.pdf. 29

Efrati, S., Catalogna, M., Abu Hamad, R., Hadanny, A., Bar-Chaim, A., Benveniste-Levkovitz, P., and Levtzion-Korach, O. (2021). Safety and humoral responses to BNT162b2 mRNA vaccination of SARS-CoV-2 previously infected and naive populations. Sci. Rep. 11, 16543.

Fialová, L., Petráčková, M., and Kuchař, O. (2017). Comparison of different enzyme-linked immunosorbent assay methods for avidity determination of antiphospholipid antibodies. J. Clin. Lab. Anal. 31, e22121.

Fisman, D.N., and Tuite, A.R. (2021). Progressive Increase in Virulence of Novel SARS-CoV-2 Variants in Ontario, Canada. medRxiv 2021.07.05.21260050.

Fraley, E., LeMaster, C., Banerjee, D., Khanal, S., Selvarangan, R., and Bradley, T. (2021). Cross-reactive antibody immunity against SARS-CoV-2 in children and adults. Cell. Mol. Immunol. 18, 1826–1828.

Gaebler, C., Wang, Z., Lorenzi, J.C.C., Muecksch, F., Finkin, S., Tokuyama, M., Cho, A., Jankovic, M., Schaefer-Babajew, D., Oliveira, T.Y., et al. (2021). Evolution of antibody immunity to SARS-CoV-2. Nature.

Gilbert, P.B., Montefiori, D.C., McDermott, A., Fong, Y., Benkeser, D., Deng, W., Zhou, H., Houchens, C.R., Martins, K., Jayashankar, L., et al. (2021). Immune Correlates Analysis of the mRNA-1273 COVID-19 Vaccine Efficacy Trial. medRxiv 2021.08.09.21261290.

Goel, R.R., Painter, M.M., Apostolidis, S.A., Mathew, D., Meng, W., Rosenfeld, A.M., Lundgreen, K.A., Reynaldi, A., Khoury, D.S., Pattekar, A., et al. (2021a). mRNA vaccines induce durable immune memory to SARS-CoV-2 and variants of concern. Science eabm0829.

Goel, R.R., Apostolidis, S.A., Painter, M.M., Mathew, D., Pattekar, A., Kuthuru, O., Gouma, S., Hicks, P., Meng, W., Rosenfeld, A.M., et al. (2021b). Distinct antibody and memory B cell responses in SARS-CoV-2 naïve and recovered individuals following mRNA vaccination. Sci. Immunol. 6.

Goldberg, Y., Mandel, M., Bar-On, Y.M., Bodenheimer, O., Freedman, L., Haas, E.J., Milo, R., Alroy-Preis, S., Ash, N., and Huppert, A. (2021). Waning immunity of the BNT162b2 vaccine: A nationwide study from Israel. medRxiv 2021.08.24.21262423.

Gong, S.Y., Chatterjee, D., Richard, J., Prévost, J., Tauzin, A., Gasser, R., Bo, Y., Vézina, D., Goyette, G., Gendron-Lepage, G., et al. (2021). Contribution of single mutations to selected SARS-CoV-2 emerging variants spike antigenicity. Virology 563, 134–145.

Hicks, J., Klumpp-Thomas, C., Kalish, H., Shunmugavel, A., Mehalko, J., Denson, J.-P., Snead, K.R., Drew, M., Corbett, K.S., Graham, B.S., et al. (2021). Serologic Cross-Reactivity of SARS-CoV-2 with Endemic and Seasonal Betacoronaviruses. J. Clin. Immunol. 41, 906–913.

Hoffmann, M., Müller, M.A., Drexler, J.F., Glende, J., Erdt, M., Gützkow, T., Losemann, C., Binger, T., Deng, H., Schwegmann-Weßels, C., et al. (2013). Differential sensitivity of bat cells to infection by enveloped RNA viruses: coronaviruses, paramyxoviruses, filoviruses, and influenza viruses. PloS One 8, e72942.

Hoffmann, M., Kleine-Weber, H., Schroeder, S., Krüger, N., Herrler, T., Erichsen, S., Schiergens, T.S., Herrler, G., Wu, N.-H., Nitsche, A., et al. (2020). SARS-CoV-2 Cell Entry Depends on ACE2 and TMPRSS2 and Is Blocked by a Clinically Proven Protease Inhibitor. Cell 181, 271–280.e8.

Isabel, S., Graña-Miraglia, L., Gutierrez, J.M., Bundalovic-Torma, C., Groves, H.E., Isabel, M.R., Eshaghi, A., Patel, S.N., Gubbay, J.B., Poutanen, T., et al. (2020). Evolutionary and structural analyses of SARS-CoV-2 D614G spike protein mutation now documented worldwide. Sci. Rep. 10, 14031.

Jackson, L.A., Anderson, E.J., Rouphael, N.G., Roberts, P.C., Makhene, M., Coler, R.N., McCullough, M.P., Chappell, J.D., Denison, M.R., Stevens, L.J., et al. (2020). An mRNA Vaccine against SARS-CoV-2 — Preliminary Report. N. Engl. J. Med. 383, 1920–1931.

JCVI (2021). Joint Committee on Vaccination and Immunisation (JCVI) advice on third primary dose vaccination, https://www.gov.uk/government/publications/third-primary-covid-19-vaccine-dose-for-people-who-are-immunosuppressed-jcvi-advice/joint-committee-on-vaccination-and-immunisation-jcvi-advice-on-third-primary-dose-vaccination.

Jennewein, M.F., MacCamy, A.J., Akins, N.R., Feng, J., Homad, L.J., Hurlburt, N.K., Seydoux, E., Wan, Y.-H., Stuart, A.B., Edara, V.V., et al. (2021). Isolation and characterization of cross-neutralizing coronavirus antibodies from COVID-19+ subjects. Cell Rep. 36, 109353.

Kim, W., Zhou, J.Q., Sturtz, A.J., Horvath, S.C., Schmitz, A.J., Lei, T., Kalaidina, E., Thapa, M., Soussi, W.B.A., Haile, A., et al. (2021). Germinal centre-driven maturation of B cell response to SARS-CoV-2 vaccination.

Li, W., Chen, Y., Prévost, J., Ullah, I., Lu, M., Gong, S.Y., Tauzin, A., Gasser, R., Vézina, D., Anand, S.P., et al. (2021). Structural Basis and Mode of Action for Two Broadly Neutralizing Antibodies Against SARS-CoV-2 Emerging Variants of Concern. BioRxiv Prepr. Serv. Biol. 2021.08.02.454546.

Lozano-Ojalvo, D., Camara, C., Lopez-Granados, E., Nozal, P., Del Pino-Molina, L., Bravo-Gallego, L.Y., Paz-Artal, E., Pion, M., Correa-Rocha, R., Ortiz, A., et al. (2021). Differential effects of the second SARS-CoV-2 mRNA vaccine dose on T cell immunity in naive and COVID-19 recovered individuals. Cell Rep. 109570.

Meulen, J. ter, Brink, E.N. van den, Poon, L.L.M., Marissen, W.E., Leung, C.S.W., Cox, F., Cheung, C.Y., Bakker, A.Q., Bogaards, J.A., Deventer, E. van, et al. (2006). Human Monoclonal Antibody Combination against SARS Coronavirus: Synergy and Coverage of Escape Mutants. PLOS Med. 3, e237.

Moore, J.P., and Klasse, P.J. (2020). COVID-19 Vaccines: “Warp Speed” Needs Mind Melds, Not Warped Minds. J. Virol. 94.

Muruato, A.E., Fontes-Garfias, C.R., Ren, P., Garcia-Blanco, M.A., Menachery, V.D., Xie, X., and Shi, P.-Y. (2020). A high-throughput neutralizing antibody assay for COVID-19 diagnosis and vaccine evaluation. Nat. Commun. 11, 4059.

Ng, K.W., Faulkner, N., Cornish, G.H., Rosa, A., Harvey, R., Hussain, S., Ulferts, R., Earl, C., Wrobel, A.G., Benton, D.J., et al. (2020). Preexisting and de novo humoral immunity to SARS-CoV-2 in humans. Science 370, 1339–1343.

Painter, M.M., Mathew, D., Goel, R.R., Apostolidis, S.A., Pattekar, A., Kuthuru, O., Baxter, A.E., Herati, R.S., Oldridge, D.A., Gouma, S., et al. (2021). Rapid induction of antigen-specific CD4+ T cells is associated with coordinated humoral and cellular immune responses to SARS-CoV-2 mRNA vaccination. Immunity S1074761321003083.

Park, J.-E., Li, K., Barlan, A., Fehr, A.R., Perlman, S., McCray, P.B., and Gallagher, T. (2016). Proteolytic processing of Middle East respiratory syndrome coronavirus spikes expands virus tropism. Proc. Natl. Acad. Sci. 113, 12262–12267.

Parry, H., Bruton, R., Stephens, C., Brown, K., Amirthalingam, G., Otter, A., Hallis, B., Zuo, J., and Moss, P. (2021). Differential immunogenicity of BNT162b2 or ChAdOx1 vaccines after extended-interval homologous dual vaccination in older people. Immun. Ageing A 18, 34.

Payne, R.P., Longet, S., Austin, J.A., Skelly, D.T., Dejnirattisai, W., Adele, S., Meardon, N., Faustini, S., Al-Taei, S., Moore, S.C., et al. (2021). Immunogenicity of standard and extended dosing intervals of BNT162b2 mRNA vaccine. Cell 0.

Pearson, C.A.B., Russell, T.W., Davies, N.G., Kucharski, A.J., CMMID COVID-19 working group, Edmunds, W.J., and Eggo, R.M. (2021). Estimates of severity and transmissibility of novel SARS-CoV-2 variant 501Y.V2 in South Africa, https://cmmid.github.io/topics/covid19/sa-novel-variant.html.

Pilishvili, T. (2021). Interim Estimates of Vaccine Effectiveness of Pfizer-BioNTech and Moderna COVID-19 Vaccines Among Health Care Personnel — 33 U.S. Sites, January–March 2021. MMWR Morb. Mortal. Wkly. Rep. 70.

Planas, D., Veyer, D., Baidaliuk, A., Staropoli, I., Guivel-Benhassine, F., Rajah, M.M., Planchais, C., Porrot, F., Robillard, N., Puech, J., et al. (2021a). Reduced sensitivity of SARS-CoV-2 variant Delta to antibody neutralization. Nature 596, 276–280.

Planas, D., Bruel, T., Grzelak, L., Guivel-Benhassine, F., Staropoli, I., Porrot, F., Planchais, C., Buchrieser, J., Rajah, M.M., Bishop, E., et al. (2021b). Sensitivity of infectious SARS-CoV-2 B.1.1.7 and B.1.351 variants to neutralizing antibodies. Nat. Med. 27, 917–924.

Polack, F.P., Thomas, S.J., Kitchin, N., Absalon, J., Gurtman, A., Lockhart, S., Perez, J.L., Pérez Marc, G., Moreira, E.D., Zerbini, C., et al. (2020). Safety and Efficacy of the BNT162b2 mRNA Covid-19 Vaccine. N. Engl. J. Med. 383, 2603–2615.

Prévost, J., and Finzi, A. (2021). The great escape? SARS-CoV-2 variants evading neutralizing responses. Cell Host Microbe 29, 322–324.

Prévost, J., Gasser, R., Beaudoin-Bussières, G., Richard, J., Duerr, R., Laumaea, A., Anand, S.P., Goyette, G., Benlarbi, M., Ding, S., et al. (2020). Cross-Sectional Evaluation of Humoral Responses against SARS-CoV-2 Spike. Cell Rep. Med. 1, 100126.

Prévost, J., Richard, J., Gasser, R., Ding, S., Fage, C., Anand, S.P., Adam, D., Vergara, N.G., Tauzin, A., Benlarbi, M., et al. (2021). Impact of temperature on the affinity of SARS-CoV-2 Spike glycoprotein for host ACE2. J. Biol. Chem. 101151.

Puranik, A., Lenehan, P.J., Silvert, E., Niesen, M.J.M., Corchado-Garcia, J., O’Horo, J.C., Virk, A., Swift, M.D., Halamka, J., Badley, A.D., et al. (2021). Comparison of two highly-effective mRNA vaccines for COVID-19 during periods of Alpha and Delta variant prevalence. R R: a language and environment for statistical computing. https://www.gbif.org/fr/tool/81287/r-a-language-and-environment-for-statistical-computing.

R studio RStudio | Open source & professional software for data science teams. https://rstudio.com/.

Rabaan, A.A., Al-Ahmed, S.H., Haque, S., Sah, R., Tiwari, R., Malik, Y.S., Dhama, K., Yatoo, M.I., Bonilla-Aldana, D.K., and Rodriguez-Morales, A.J. (2020). SARS-CoV-2, SARS-CoV, and MERS-COV: A comparative overview. Infez. Med. 28, 174–184.

Rambaut, A., Loman, N., Pybus, O., Barclay, W., Barrett, J., Carabelli, A., Connor, T., Peacock, T., Robertson, D.L., and Volz, E. (2020). Preliminary genomic characterisation of an emergent SARS-CoV-2 lineage in the UK defined by a novel set of spike mutations - SARS-CoV-2 coronavirus / nCoV-2019 Genomic Epidemiology.

Rudnick, S.I., and Adams, G.P. (2009). Affinity and avidity in antibody-based tumor targeting. Cancer Biother. Radiopharm. 24, 155–161.

Sabino, E.C., Buss, L.F., Carvalho, M.P.S., Prete, C.A., Crispim, M.A.E., Fraiji, N.A., Pereira, R.H.M., Parag, K.V., Peixoto, P. da S., Kraemer, M.U.G., et al. (2021). Resurgence of COVID-19 in Manaus, Brazil, despite high seroprevalence. The Lancet 397, 452–455.

Sahin, U., Muik, A., Derhovanessian, E., Vogler, I., Kranz, L.M., Vormehr, M., Baum, A., Pascal, K., Quandt, J., Maurus, D., et al. (2020). COVID-19 vaccine BNT162b1 elicits human antibody and T H 1 T cell responses. Nature 586, 594–599.

Sarkar, J.P., Saha, I., Seal, A., Maity, D., and Maulik, U. (2021). Topological Analysis for Sequence Variability: Case Study on more than 2K SARS-CoV-2 sequences of COVID-19 infected 54 countries in comparison with SARS-CoV-1 and MERS-CoV. Infect. Genet. Evol. 88, 104708.

Skowronski, D., and De Serres, G. (2021). Safety and Efficacy of the BNT162b2 mRNA Covid-19 Vaccine. N. Engl. J. Med. NEJMc2036242.

Skowronski, D.M., Setayeshgar, S., Febriani, Y., Ouakki, M., Zou, M., Talbot, D., Prystajecky, N., Tyson, J.R., Gilca, R., Brousseau, N., et al. (2021). Two-dose SARS-CoV-2 vaccine effectiveness with mixed schedules and extended dosing intervals: test-negative design studies from British Columbia and Quebec, Canada (Infectious Diseases (except HIV/AIDS)).

Stamatatos, L., Czartoski, J., Wan, Y.-H., Homad, L.J., Rubin, V., Glantz, H., Neradilek, M., Seydoux, E., Jennewein, M.F., MacCamy, A.J., et al. (2021). mRNA vaccination boosts cross-variant neutralizing antibodies elicited by SARS-CoV-2 infection. Science eabg9175.

Tang, J.W., Toovey, O.T.R., Harvey, K.N., and Hui, D.S.C. (2021). Introduction of the South African SARS-CoV-2 variant 501Y.V2 into the UK. J. Infect. 82, e8–e10.

Tartof, S.Y., Slezak, J.M., Fischer, H., Hong, V., Ackerson, B.K., Ranasinghe, O.N., Frankland, T.B., Ogun, O.A., Zamparo, J.M., Gray, S., et al. (2021). Six-Month Effectiveness of BNT162B2 mRNA COVID-19 Vaccine in a Large US Integrated Health System: A Retrospective Cohort Study (Rochester, NY: Social Science Research Network).

Tauzin, A., Nayrac, M., Benlarbi, M., Gong, S.Y., Gasser, R., Beaudoin-Bussières, G., Brassard, N., Laumaea, A., Vézina, D., Prévost, J., et al. (2021). A single dose of the SARS-CoV-2 vaccine BNT162b2 elicits Fc-mediated antibody effector functions and T cell responses. Cell Host Microbe 0.

Tegally, H., Wilkinson, E., Giovanetti, M., Iranzadeh, A., Fonseca, V., Giandhari, J., Doolabh, D., Pillay, S., San, E.J., Msomi, N., et al. (2020). Emergence and rapid spread of a new severe acute respiratory syndrome-related coronavirus 2 (SARS-CoV-2) lineage with multiple spike mutations in South Africa. MedRxiv 2020.12.21.20248640.

Ullah, I., Prévost, J., Ladinsky, M.S., Stone, H., Lu, M., Anand, S.P., Beaudoin-Bussières, G., Symmes, K., Benlarbi, M., Ding, S., et al. (2021). Live imaging of SARS-CoV-2 infection in mice reveals that neutralizing antibodies require Fc function for optimal efficacy. Immunity S1074-7613(21)00347-2.

Urbanowicz, R.A., Tsoleridis, T., Jackson, H.J., Cusin, L., Duncan, J.D., Chappell, J.G., Tarr, A.W., Nightingale, J., Norrish, A.R., Ikram, A., et al. (2021). Two doses of the SARS-CoV-2 BNT162b2 vaccine enhances antibody responses to variants in individuals with prior SARS-CoV-2 infection. Sci. Transl. Med. eabj0847.

Volz, E., Mishra, S., Chand, M., Barrett, J.C., Johnson, R., Geidelberg, L., Hinsley, W.R., Laydon, D.J., Dabrera, G., O’Toole, Á., et al. (2021). Transmission of SARS-CoV-2 Lineage B.1.1.7 in England: Insights from linking epidemiological and genetic data. MedRxiv 2020.12.30.20249034.

Wall, E.C., Wu, M., Harvey, R., Kelly, G., Warchal, S., Sawyer, C., Daniels, R., Hobson, P., Hatipoglu, E., Ngai, Y., et al. (2021). Neutralising antibody activity against SARS-CoV-2 VOCs B.1.617.2 and B.1.351 by BNT162b2 vaccination. The Lancet 397, 2331–2333.

Walls, A.C., Park, Y.-J., Tortorici, M.A., Wall, A., McGuire, A.T., and Veesler, D. (2020). Structure, Function, and Antigenicity of the SARS-CoV-2 Spike Glycoprotein. Cell 181, 281-292.e6.

Wang, P., Nair, M.S., Liu, L., Iketani, S., Luo, Y., Guo, Y., Wang, M., Yu, J., Zhang, B., Kwong, P.D., et al. (2021a). Antibody Resistance of SARS-CoV-2 Variants B.1.351 and B.1.1.7.

Wang, Q., Lei, Y., Lu, X., Wang, G., Du, Q., Guo, X., Xing, Y., Zhang, G., and Wang, D. (2019). Urea-mediated dissociation alleviate the false-positive Treponema pallidum-specific antibodies detected by ELISA. PLOS ONE 14, e0212893.

Wang, Z., Muecksch, F., Schaefer-Babajew, D., Finkin, S., Viant, C., Gaebler, C., Hoffmann, H.-H., Barnes, C.O., Cipolla, M., Ramos, V., et al. (2021b). Naturally enhanced neutralizing breadth against SARS-CoV-2 one year after infection. Nature 595, 426–431.

WHO (2021). Interim recommendations for use of the Pfizer–BioNTech COVID-19 vaccine, BNT162b2, under Emergency Use Listing, https://www.who.int/publications/i/item/WHO-2019-nCoV-vaccines-SAGE_recommendation-BNT162b2-2021.1.

World Health Organization WHO Coronavirus (COVID-19) Dashboard. https://covid19.who.int.

